# Meta-analysis of CYP2C19 and CYP2D6 metabolic activity on antidepressant response from 13 clinical studies using genotype imputation

**DOI:** 10.1101/2023.06.26.23291890

**Authors:** Danyang Li, Oliver Pain, Chiara Fabbri, Win Lee Edwin Wong, Chris Wai Hang Lo, Stephan Ripke, Annamaria Cattaneo, Daniel Souery, Mojca Z. Dernovsek, Neven Henigsberg, Joanna Hauser, Glyn Lewis, Ole Mors, Nader Perroud, Marcella Rietschel, Rudolf Uher, Wolfgang Maier, Bernhard T. Baune, Joanna M. Biernacka, Guido Bondolfi, Katharina Domschke, Masaki Kato, Yu-Li Liu, Alessandro Serretti, Shih-Jen Tsai, Richard Weinshilboum, the GSRD Consortium, the Major Depressive Disorder Working Group of the Psychiatric Genomics Consortium, Andrew M. McIntosh, Cathryn M. Lewis

**Affiliations:** Social, Genetic and Developmental Psychiatry Centre, King’s College London, London, GB; Maurice Wohl Clinical Neuroscience Institute, Department of Basic and Clinical Neuroscience, Institute of Psychiatry, Psychology and Neuroscience, King’s College London, London, GB; Department of Biomedical and Neuromotor Sciences, University of Bologna, Bologna, IT; Department of Pharmacology, Yong Loo Lin School of Medicine, National University of Singapore, Singapore, SG; Department of Psychiatry and Psychotherapy, Universitätsmedizin Berlin Campus Charité Mitte, Berlin, DE; Analytic and Translational Genetics Unit, Massachusetts General Hospital, Boston, MA, US; Biological Psychiatry Laboratory, IRCCS Fatebenefratelli, Brescia, IT; Department of Pharmacological and Biomedical Sciences, University of Milan, Milan, IT; Laboratoire de Psychologie Medicale, Universitè Libre de Bruxelles and Psy Pluriel, Centre Européen de Psychologie Medicale, Brussels, BE; University Psychiatric Clinic, University of Ljubliana, Ljubljana, SI; Department of Psychiatry, Croatian Institute for Brain Research, University of Zagreb Medical School, Zagreb, HR; Psychiatric Genetic Unit,, Poznan University of Medical Sciences, Poznan, PL; Division of Psychiatry, University College London, London, GB; Psychosis Research Unit, Aarhus University Hospital - Psychiatry, Aarhus, DK; Department of Psychiatry, Geneva University Hospitals, Geneva, CH; Department of Genetic Epidemiology in Psychiatry, Medical Faculty Mannheim, University of Heidelberg, Central Institute of Mental Health, Mannheim, DE; Department of Psychiatry, Dalhousie University, Halifax, NS, CA; Department of Psychiatry and Psychotherapy, University of Bonn, Bonn, DE; Department of Psychiatry, University of Münster, Münster, DE; Florey Institute for Neuroscience and Mental Health, University of Melbourne, Melbourne, AU; Department of Psychiatry, Melbourne Medical School, University of Melbourne, Melbourne, AU; Department of Psychiatry and Psychology, Mayo Clinic, Rochester, MN, USA; Department of Quantitative Health Sciences, Mayo Clinic, Rochester, MN, USA; Department of Psychiatry and Psychotherapy, Medical Center, University of Freiburg, Freiburg, DE; Department of Neuropsychiatry, Kansai Medical University, Osaka, JP; Center for Neuropsychiatric Research, National Health Research Institutes, Miaoli, TW; Department of Psychiatry, Taipei Veterans General Hospital, Taipei, TW; Division of Psychiatry, School of Medicine, National Yang-Ming University, Taipei, TW; Department of Molecular Pharmacology and Experimental Therapeutics, Mayo Clinic, Rochester, MN, USA; Division of Psychiatry, University of Edinburgh, Edinburgh, GB; Department of Medical & Molecular Genetics, King’s College London, London, GB

**Keywords:** antidepressants, pharmacogenetics, CYP2C19, CYP2D6

## Abstract

Cytochrome P450 enzymes including CYP2C19 and CYP2D6 are important for antidepressant metabolism and polymorphisms of these genes have been determined to predict metabolite levels. Nonetheless, more evidence is needed to understand the impact of genetic variations on antidepressant response. In this study, individual clinical and genetic data from 13 studies of European and East Asian ancestry populations were collected. The antidepressant response was clinically assessed as remission and percentage improvement. Imputed genotype was used to translate genetic polymorphisms to metabolic phenotypes (poor, intermediate, normal, and rapid+ultrarapid) of CYP2C19 and CYP2D6. The association of CYP2C19 and CYP2D6 metabolic phenotypes with treatment response was examined using normal metabolizers as the reference. Among 5843 depression patients, a higher remission rate was found in CYP2C19 poor metabolizers compared to normal metabolizers at nominal significance but did not survive after multiple testing correction (OR=1.46, 95% CI [1.03, 2.06], p=0.033, heterogeneity I^2^=0%, subgroup difference p=0.72). No metabolic phenotype was associated with percentage improvement from baseline. After stratifying by antidepressants primarily metabolized by CYP2C19 and CYP2D6, no association was found between metabolic phenotypes and antidepressant response. Metabolic phenotypes showed differences in frequency, but not effect, between European- and East Asian-ancestry studies. In conclusion, metabolic phenotypes imputed from genetic variants using genotype were not associated with antidepressant response. CYP2C19 poor metabolizers could potentially contribute to antidepressant efficacy with more evidence needed. CYP2D6 structural variants cannot be imputed from genotype data, limiting inference of pharmacogenetic effects. Sequencing and targeted pharmacogenetic testing, alongside information on side effects, antidepressant dosage, depression measures, and diverse ancestry studies, would more fully capture the influence of metabolic phenotypes.

## Introduction

Antidepressants are the first-line treatment for moderate or severe depression, however efficacy varies, and side effects are common ^1^. Approximately 35% of patients reach remission after treatment with a single antidepressant but a substantial proportion require further treatment, with many developing treatment-resistant depression^2–5^. Even within the same antidepressant class, treatment responses vary substantially. For example, selective serotonin reuptake inhibitors (SSRIs), the most widely prescribed antidepressants, could lead to remission in 30-45% of patients ^6^. Differences in response rate may be due to many factors including drug-drug interactions ^7^, depression subtypes ^8,9^, comorbidity ^10^, smoking ^11^, and genetic variation, particularly in drug metabolism genes.

Pharmacogenetics utilizes genetic variation that plays a role in medication action and metabolism to facilitate individualized prescription, thus improving the treatment efficacy, and reducing undesirable effects ^12^. In antidepressants, current evidence and prescribing guidelines support cytochrome P450 (CYP) genes for pharmacogenetic testing, in which *CYP2C19* and *CYP2D6* have been widely examined for drug efficacy and side effects ^12–15^. Both *CYP2C19* and *CYP2D6* are highly polymorphic, with genetic haplotypes defined by the star allele nomenclature ^16^. These star alleles can be classified into different metabolic phenotypes, such as poor metabolizers (PMs), intermediate metabolizers (IMs), normal metabolizers (NMs), rapid and ultrarapid metabolizers (RMs/UMs) according to Clinical Pharmacogenetics Implementation Consortium (CPIC) guidelines ^14,15^. Compared to NMs, PMs and IMs have an increased risk of adverse effects because of a lower metabolism rate and elevated drug serum concentrations, which may also increase treatment efficacy. RMs and UMs, on the other hand, facilitate the metabolic process to reduce drug exposure and may lead to treatment failure through a lack of efficacy.

Clinical studies have shown that genetic variation in these metabolizing enzymes is clearly associated with metabolite levels, but the link between genetic variation and treatment response or side effects is more complicated. For example, in the GENDEP study, CYP2C19 and CYP2D6 genotypes were associated with serum concentration of escitalopram and nortriptyline, but did not predict treatment response ^17^. A meta-analysis of 94 studies assessed the relationship between psychiatric drug exposure (dose-normalized plasma level) and metabolising status of CYP2C19 and CYP2D6, observing exposure differences in escitalopram and sertraline ^18^. However, treatment effectiveness of these antidepressants was not associated with CYP2C19 genotypes in a large retrospective study based on participant self-report ^19^.

Guidelines have been developed for antidepressant use based on CYP2D6 and CYP2C19 metabolizing status. For instance, CPIC guidelines for CYP2D6 suggest a 50% dose reduction of vortioxetine, paroxetine and most tricyclic antidepressants for PMs, and alternative antidepressants that are not predominantly metabolized by CYP2D6 are advised for UMs ^14,15^. However, evidence is still accruing to confirm the role of pharmacogenetic testing to guide antidepressant prescribing ^12^.

Both CYP2C19 and CYP2D6 require in-depth assessment of variation to fully determine star alleles and metabolizer status but it is complicated by structural variants and pseudogenes, particularly for CYP2D6. Full assessment cannot be achieved through genotyping but requires pharmacogenetic-specific tests (e.g., targeted arrays, sequencing) ^20–22^. However, many research studies have genome-wide genotyping and lack full pharmacogenetic assessment. In this study, we used imputation from genome-wide genotyping to estimate the metabolic status and test for association of CYP2C19 and CYP2D6 metabolic phenotypes with clinically evaluated treatment response across multiple antidepressants. We combined clinical and genetic data from 13 clinical studies, with 5843 participants, of European and East Asian ancestry. We investigated whether genotype-determined PMs, IMs, and RMs/UMs of CYP2C19 and CYP2D6 showed differential antidepressant efficacy, compared to NMs. This unique resource provides additional evidence of the relationship between CYP gene metabolic phenotypes and treatment response, and may further determine whether genotype-determined metabolizer status could add useful information for individualized prescribing of antidepressants.

## Methods

### Samples

The clinical studies analysed have been described in detail previously ^3^. In brief, 10 studies with European ancestry and 3 studies from East Asia were included. All participants had a diagnosis of major depressive disorder (MDD) and received at least one antidepressant, with treatment response collected at baseline, and for 4-12 weeks post-baseline. Informed consent was obtained from all participants. We assessed two antidepressant response outcomes of remission and percentage improvement. Remission was a binary outcome defined as a reduction of the depression symptoms to a prespecified criteria of the rating scale. Percentage improvement was a continuous measure calculated from the proportional decrease (or increase) of depression symptom score from baseline. The percentage improvement was standardized (mean 0, standard deviation 1) within study to allow comparability of different scales across the studies (e.g., HAMD (Hamilton Depression Rating Scale), MADRS (Montgomery Åsberg Depression Rating Scale), QIDSC (Quick Inventory of Depressive Symptomatology)). Demographic and clinical variables of age, sex, MDD baseline severity and antidepressant prescription information were available in each study (Supplementary Table 1).

Detailed procedures of genotyping have been reported elsewhere ^23–31^. Quality control and imputation were processed using the standard ‘RICOPILI’ pipeline from the Psychiatric Genomics Consortium (PGC) with 1000 Genomes Project multi-ancestry reference panel ^32^. Each step was performed separately in European and East Asian ancestry studies following standard PGC protocols. Study details can be found in Supplementary Table 1 and the previous study ^3^.

### Star alleles and metabolic phenotypes

Using best guess imputed genotype calls, phasing was conducted separately on the genetic regions of *CYP2C19* and *CYP2D6* obtained from PharmGKB (https://www.pharmgkb.org/). The haplotype was determined in each sample using SHAPEIT4 software and the 1000 Genomes Project multi-ancestry reference panel ^33^. To fully utilize phased SNPs and translate them to star alleles, we first extracted all SNPs used to define *CYP2C19* and *CYP2D6* star alleles from the CPIC definition tables (https://cpicpgx.org/; downloaded June 2022). These SNPs were then matched to the phased data, and matching SNPs were assigned to star alleles following the CPIC guidelines. If a star allele was defined by more than one SNP, it was counted only when all the defined SNPs were observed. Each star allele was annotated as having no, decreased, normal, or increased function with corresponding activity value based on CPIC definition tables and the previous literature (Supplementary Table 2) ^34^. The reference allele (*1) was assigned to haplotypes that had no annotated functional star alleles or had uncertain or unknown functional alleles of *CYP2D6*. Because structural variants cannot be determined from genotype data, CYP2D6 rapid and ultrarapid metabolizers were not included. Next, we calculated the activity score for each individual by adding the activity values of the two star alleles. Metabolic phenotypes (PM, IM, NM) of CYPC19 were classified based on CPIC and CYP2C19 rapid+ultrarapid metabolizers were defined as individuals carrying at least one increased functional allele (*17) ^15^. CYP2D6 phenotypes (PM, IM, NM) were determined following consensus recommendations from the CPIC and the Dutch Pharmacogenetics Working Group (DPWG) ^35^. To validate the defined metabolic phenotypes, we compared phenotype concordance with that previously derived in the GENDEP using Roche AmpliChip CYP450 microarray and TaqMan SNP genotyping ^17^. After harmonizing the metabolizer status, the concordance rate (percentage of individuals assigned the same metabolic phenotypes) was 96.4% for CYP2C19 and 79.9% for CYP2D6 (Supplementary Table 3).

### Statistical analyses

#### Associations

We used the NMs in CYP2C19 and CYP2D6 as the reference group to examine the effect of other metabolizer groups on antidepressant response. For remission, logistic regression was used to evaluate the association with CYP2C19 and CYP2D6 metabolic phenotypes in each study, including age, sex, and MDD baseline severity as covariates. For percentage improvement, linear regression with CYP2C19 and CYP2D6 metabolic phenotypes, adjusting for age and sex, was used to test for association with metabolic phenotypes. The correlation between MDD baseline score and percentage improvement was very low (Pearson correlation = 0.042), so we did not add MDD baseline severity as a covariate. We next stratified into ‘antidepressant groups’, with drugs that were primarily metabolized by either CYP2C19 or CYP2D6, based on the clinical annotation of Level 1A in PharmGKB ^13,36^(Supplementary Table 4). Stratifying participants by CYP2C19- and CYP2D6-metabolized antidepressants, we repeated the analyses of remission and percentage improvement in 10 studies with CYP2C19-metabolized antidepressants (3390 participants) and 6 studies with CYP2D6-metabolized antidepressants (1223 participants) (Supplementary Figure 1).

#### Meta-analyses

In each study, odds ratios (ORs) of remission, and Standard Mean Differences (SMDs, Cohen’s D) of percentage improvement, with standard errors of both effect sizes, for each metabolizer group were extracted. We applied random effect meta-analysis since the true effects were assumed to be heterogeneous due to the difference in factors such as study populations, antidepressants prescribed, and outcome measurements. The effect sizes in each study were pooled, and inverse-variance weighted. The between-study heterogeneity was quantified by *I*^*2*^ statistic and heterogeneity variance *τ*^*2*^ using the Paule-Mandel method for ORs and restricted maximum-likelihood estimator for SMDs. The significance was tested by Cochran’s Q at p < 0.05. Additionally, subgroup meta-analyses were applied to test the hypothesis that effects differed between European and East Asian ancestry. We assumed both ancestries shared a common between-study heterogeneity (*τ*^*2*^) due to a small number of studies from East Asia. Cochran’s Q was used to determine whether the differences between subgroups could be explained by true effect differences or by sampling errors alone. We performed meta-analyses in all samples for CYP2C19 and CYP2D6 metabolic phenotypes and then stratified the analyses by antidepressant groups for the corresponding metabolizer effects. We used p value < 0.05 as nominal significance, and corrected for multiple testing for the 5 independent tests of metabolic phenotypes compared with NMs (3 phenotypes in CYP2C19 and 2 phenotypes in CYP2D6), giving a Bonferroni corrected p value of 0.01 (0.05/5). No correction across outcomes (remission and percentage improvement) was applied, due to their high correlations. All meta-analyses were performed by ‘meta’ package in R 4.2.1.

The power of the meta-analysis was calculated by ‘dmetar’ package in R 4.2.1. Using the sample size of PMs (N = 179) and NMs (N = 2289) in CYP2C19, the meta-analysis had over 80% power to detect SMD of 0.074 and OR 1.15 with no effect heterogeneity, or SMD 0.085 and OR 1.17 with low heterogeneity, at a significance level p = 0.01.

### Sensitivity tests

Four sensitivity analyses were performed. Firstly, each participant’s activity score was calculated as a continuous measure to assess metabolic activity and compared to the metabolizer groups. We tested CYP2C19 and CYP2D6 metabolic effects represented by activity scores using the same analyses described above. For the percentage improvement outcome, correlations were assessed between activity scores and residuals of percentage improvement after regressing out age and sex, and restricted maximum-likelihood estimator was used to estimate between-study heterogeneity of correlations in the meta-analyses. Secondly, the impact of baseline depression severity on percentage improvement was assessed by including it as a covariate in the linear regression analyses. Thirdly, to test how small studies might be impacting results, we reran the meta-analysis of CYP2C19 PM on the remission outcome including only studies with at least 10 PMs present. Finally, the association of CYP2C19 metabolic phenotype with citalopram and escitalopram were measured to compare the effect with all samples and CYP2C19 antidepressant group.

## Results

### Characteristics of star alleles and metabolic phenotypes

Seven star alleles in *CYP2C19* and 16 alleles in *CYP2D6* were identified from the imputed genotype data and were classified as having no, decreased, normal and increased function (Supplementary Table 2). In general, alleles had similar frequencies in studies of the same ancestry group (Supplementary Figure 2). The reference alleles (*1) were the most common, with mean frequency 62.8% in *CYP2C19*, and 39.2% in *CYP2D6* in European ancestry studies, and frequencies of 62.1% and 34.2% in East Asian studies. Other high frequency alleles in European-ancestry studies were *17 (22.0%) in *CYP2C19* and *4 (19.8%) in *CYP2D6*, while *CYP2C19* *2 (30.5%) and *CYP2D6* *10 (48.6%) had high frequencies in East Asian studies. A total of 5843 individuals with remission or percentage improvement outcome in 13 studies were analysed. Four metabolizer groups (PMs, IMs, NMs, RMs+UMs) for CYP2C19 and three metabolizer groups (PMs, IMs, and NMs) for CYP2D6 were translated from star alleles. In both genes, the most common metabolizer group was NMs, and the rarest was PMs (Table 1). Compared with the East Asians, the European-ancestry studies had a lower proportion of CYP2C19 PMs and IMs, and higher proportion of RMs+UMs. CYP2D6 PMs were only found in the European-ancestry studies (Figure 1, differences between ancestries, Wilcoxon test: CYP2C19 PMs p = 0.007, CYP2C19 IMs p = 0.007, CYP2C19 RMs+UMs p = 0.007, CYP2D6 PMs p = 0.014). For the 12 antidepressants metabolized primarily by either CYP2C19 or CYP2D6, the same distribution of metabolic phenotypes was found in both antidepressant groups (Supplementary Table 5).

**Figure 1.**
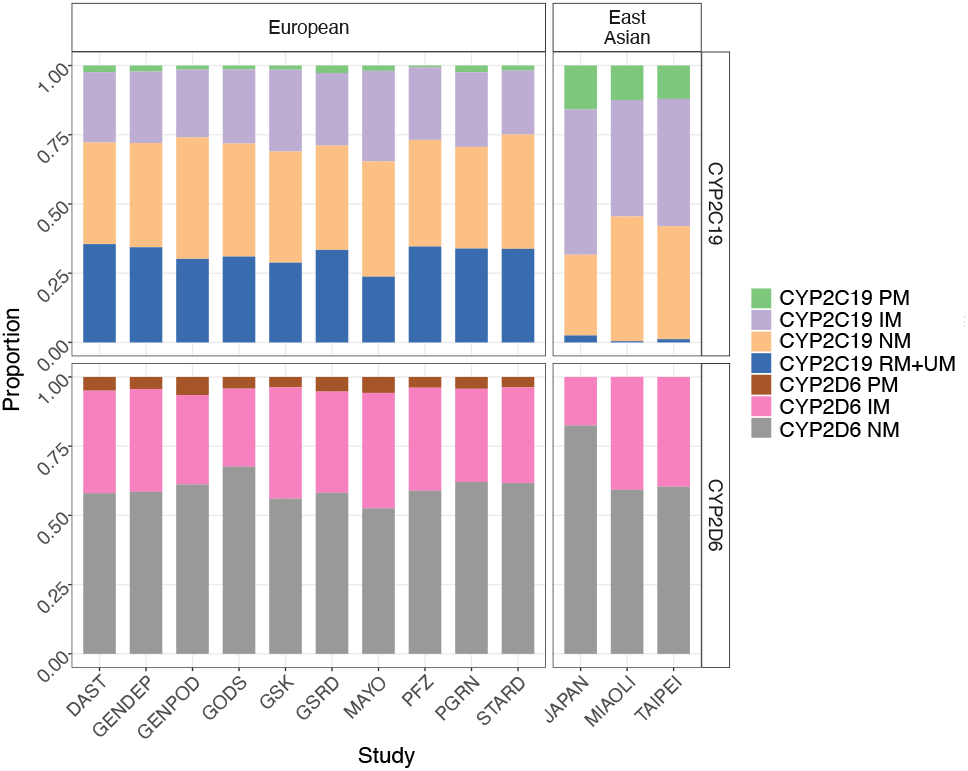
Proportion of metabolic phenotypes in each cohort. DAST: Depression and Sequence of Treatment, GENDEP: Genome Based Therapeutic Drugs for Depression, GENPOD: GENetic and clinical Predictors Of treatment response in Depression, GODS: Geneva Outpatient Depression Study, GSK: Glaxo Smith Kline, GSRD: Group for the Study of Resistant Depression, PFZ: Pfizer, PGRN: Pharmacogenomics Research Network Antidepressant Medication Pharmacogenomic Study, STARD, Sequenced Treatment Alternatives to Relieve Depression. PM: poor metabolizer, IM: intermediate metabolizer, NM: normal metabolizer, RM+UM: rapid+ultrarapid metabolizer

**Table 1.** Sample characteristics.

|  | **CYP2C19** |  |  |  | **CYP2D6** |  |  |
| --- | --- | --- | --- | --- | --- | --- | --- |
|  | **PM**<br>**(N=179)** | **IM**<br>**(N=1601)** | **NM**<br>**(N=2289)** | **RM+UM**<br>**(N=1774)** | **PM**<br>**(N=249)** | **IM**<br>**(N=2087)** | **NM**<br>**(N=3507)** |
| Remission | 89 (49.7%) | 607 (37.9%) | 885 (38.7%) | 662 (37.3%) | 96 (38.6%) | 796 (38.1%) | 1351 (38.5%) |
| Percentage Improvement | 0.12 (1.10) | 0.00 (0.98) | -0.01 (1.01) | 0.00 (0.99) | -0.02 (0.98) | 0.01 (0.98) | 0.00 (1.00) |
| Age | 45.15 (14.53) | 44.76 (14.64) | 44.44 (14.19) | 44.95 (14.17) | 43.41 (14.62) | 44.51 (14.29) | 44.91 (14.31) |
| Sex (Female) | 112 (62.6%) | 987 (61.6%) | 1438 (62.8%) | 1112 (62.7%) | 154 (61.8%) | 1301 (62.3%) | 2194 (62.6%) |
| Ancestry (European) | 110 (61.5%) | 1360 (84.9%) | 2078 (90.8%) | 1768 (99.7%) | 249 (100%) | 1902 (91.1%) | 3165 (90.2%) |
| CYP2D6 |  |  |  |  |  |  |  |
| PM | 7 (3.9%) | 55 (3.4%) | 96 (4.2%) | 91 (5.1%) | - | - | - |
| IM | 58 (32.4%) | 598 (37.4%) | 785 (34.3%) | 646 (36.4%) | - | - | - |
| NM | 114 (63.7%) | 948 (59.2%) | 1408 (61.5%) | 1037 (58.5%) | - | - | - |

Mean with standard deviation for continuous variables and frequency with proportion for categorical variables were displayed
PM: poor metabolizer, IM: intermediate metabolizer, NM: normal metabolizer, RM+UM: rapid+ultrarapid metabolizer

### Meta-analyses of CYP2C19 and CYP2D6 metabolic phenotypes in all samples

The association of metabolizer status with antidepressant response was first assessed in all samples, across all antidepressants. The remission rate and mean percentage improvement in each metabolizer group are presented in Table 1. Overall, PMs in CYP2C19 showed a higher remission rate with nominal significance (OR = 1.46, 95% CI [1.03, 2.06], p = 0.033, Figure 2a) but did not meet correction for multiple testing. The percentage improvement analysis showed a non-significant higher efficacy in PMs (SMD = 0.13, 95% CI [-0.03, 0.29], p = 0.101). Other metabolic phenotypes in CYP2C19 and CYP2D6 had no difference from NMs in both outcomes (Figure 2a). Subgroup meta-analyses found no heterogeneity in the effect of CYP2C19 PMs in all cohorts or between ancestry groups (all cohorts: I^2^ = 0%, *τ*^2^ = 0, p = 0.81; between groups: *χ*^2^ = 0.13, p = 0.72). In other metabolic phenotypes of both genes, no significant heterogeneity was detected (CYP2D6 PMs: all cohorts: I^2^ = 21%, *τ*^2^ = 0.07, p = 0.26, Supplementary Figure 3, 4).

**Figure 2.**
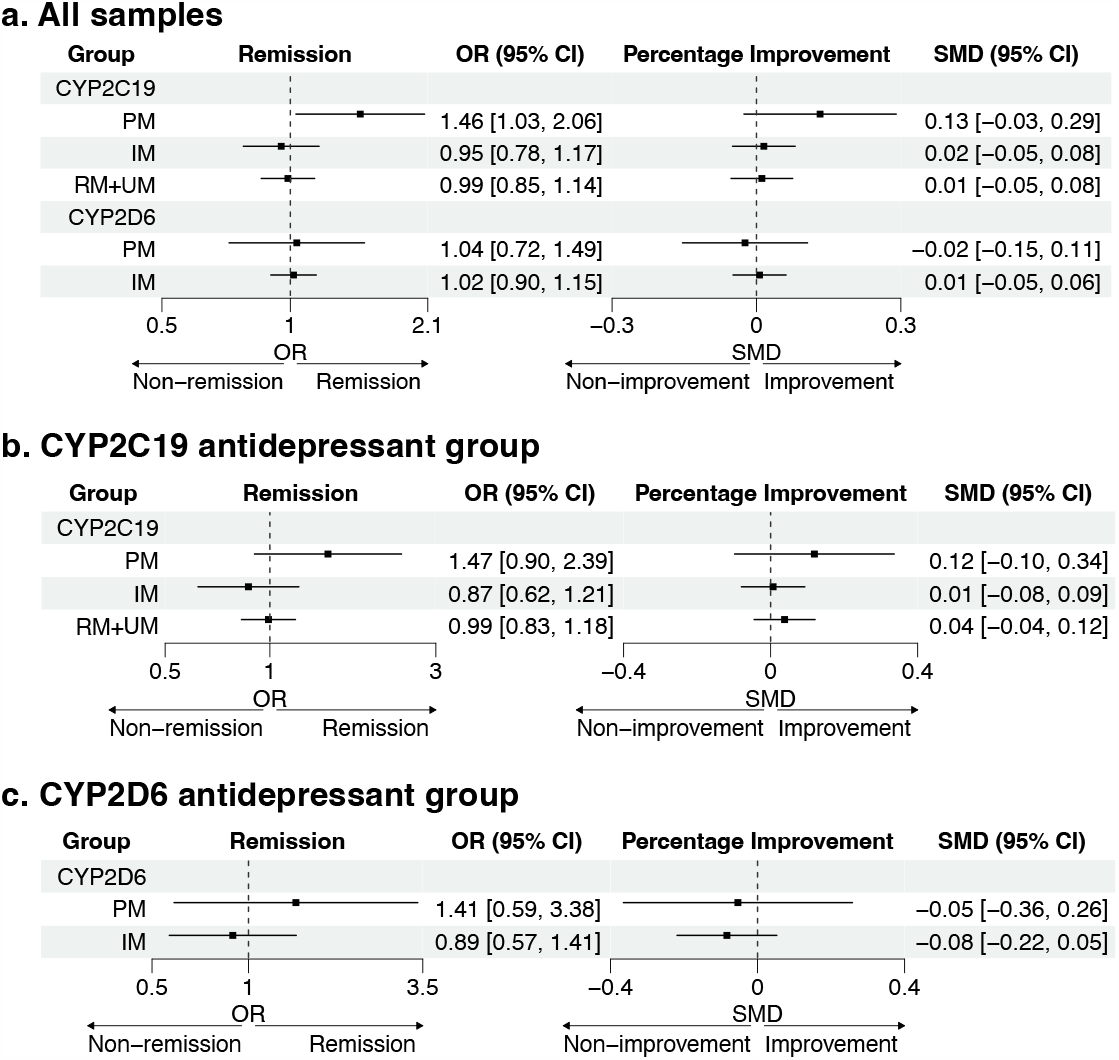
Association of CYP2C19 and CYP2D6 metabolizer status with antidepressant outcomes. PM: poor metabolizer, IM: intermediate metabolizer, RM+UM: rapid+ultrarapid metabolizer, OR: odd ratio, SMD: standard mean difference, CI: confidence interval

### Meta-analyses of CYP2C19 and CYP2D6 metabolic phenotypes stratified by antidepressant groups

Next, to determine if the metabolic activity was associated with response in antidepressants that were primarily metabolized by CYP2C19 or CYP2D6 ^13^, meta-analyses were stratified with 7 CYP2C19-metabolized antidepressants and 9 for CYP2D6 (Supplementary Table 4). CYP2C19 PMs showed a similar trend to the results in all samples, with a higher remission rate and percentage improvement compared to NMs (remission: OR = 1.47, 95% CI [0.90, 2.39], p = 0.121; percentage improvement: SMD = 0.12, 95% CI [-0.10, 0.34], p = 0.282, Figure 2b, c) but the association was not significant. Other metabolizer groups were not associated with response. Detailed results for each study can be found in Supplementary Figures 5 and 6. As a comparison, metabolic effect was tested in the antidepressant groups that were not primarily metabolized by CYP2C19 or CYP2D6. Detailed results are shown in Supplementary Figure 7.

### Sensitivity test

Finally, four sensitivity tests were performed. First, the meta-analyses were repeated using the activity score as a quantitative measurement of metabolic activity to compare the results with the primary analyses. The activity scores differed between European and East Asian studies, with Europeans having higher scores for both CYP2C19 and CYP2D6 (Wilcoxon test: CYP2C19 p = 0.007; CYP2D6 p = 0.028, Supplementary Figure 8). However, activity score was not associated with the outcomes of remission or percentage improvement (Supplementary Table 6). In the second sensitivity test, baseline severity of depression was added as an additional covariate in the analyses of percentage improvement. As in the primary analyses, PMs in CYP2C19 had higher, but non-significant SMD of percentage improvement (SMD = 0.13, 95% CI [-0.03, 0.29], p = 0.103). No clear pattern was found in tests of other metabolizers (Supplementary Table 7). Furthermore, we meta-analysed the CYP2C19 PMs for remission by including only studies with more than 10 CYP2C19 PMs. A higher rate of remission was observed in CYP2C19 PMs from 8 studies confirming the association found in the main analyses (OR = 1.56, 95% CI [1.09; 2.24], p = 0.016). Lastly, we tested the effect of CYP2C19 on citalopram and escitalopram antidepressants. No significant findings were detected, with the strongest effects found in CYP2C19 PMs, showing a non-significant increase in the remission rate and percentage improvement compared to NMs (remission: OR = 1.41, 95% CI [0.84, 2.34], p = 0.192, percentage improvement: SMD = 0.069, 95% CI [-0.16, 0.30], p = 0.559, Supplementary Table 8).

## Discussion

In this study, we leveraged 13 clinical studies (10 of European-ancestry and 3 from East Asia) to meta-analyse the association of CYP2C19 and CYP2D6 metabolic phenotypes with antidepressant response, using remission and percentage improvement as outcome measures. Using the available imputed genotype data, we identified 7 star alleles of *CYP2C19* and 16 star alleles of *CYP2D6*. We found CYP2C19 PMs had a higher remission rate compared to CYP2C19 NMs in all samples (OR = 1.46; 95% CI [1.03, 2.06]), which reached nominal significance but was not significant at the multiple testing threshold. CYP2C19 PMs also had a higher remission rate in antidepressants primarily metabolized by CYP2C19 (OR = 1.47, 95% CI [0.90, 2.39]) but differences were not significant. No difference in percentage improvement was seen between PMs and NMs. Other metabolizer groups in CYP2C19 and CYP2D6 showed no association with either remission or percentage improvement. Although there were differences in the frequency of star alleles and in the proportions of metabolic phenotypes between European and East Asian ancestry studies, the impact of metabolic phenotypes was similar.

In *CYP2C19*, our analysis pipeline detected 7 star alleles including all tier 1 alleles (*2, *3 and *17) and two tier 2 alleles (*8, *35) demonstrating a good coverage of imputed genotype for *CYP2C19* region ^37^. Nevertheless, only a moderate relationship was detected with CYP2C19 PMs with the remission outcome. Other metabolizer statuses were not associated with treatment outcomes. When testing the PMs restricted to antidepressants largely metabolized by CYP2C19, a similar effect size was detected but showed no significance, suggesting a loss of power. Other meta-analyses, retrospective studies, and clinical cohorts have replicated a higher antidepressant efficacy of CYP2C19 PMs ^19,38–40^. However, a null effect or an opposite association of CYP2C19 slow metabolizers for lower antidepressant efficacy was observed in smaller samples ^17,41,42^. This discrepancy may be due to different criteria for study participants, MDD severity, dropout rates, medication prescribed, and lack of information on other associated factors such as antidepressant dosage. Given the heterogeneity of patients and potential confounding variables, our results need further replication to understand the role of CYP2C19 metabolizers under different circumstances. In addition to treatment efficacy, PMs of CYP2C19 were also associated with worse antidepressant tolerability, although these features were not assessed in our study ^19,39^. CPIC and the Dutch Pharmacogenetics Working Group (DPWG) have recommended reducing the starting dose of escitalopram, citalopram, and sertraline for CYP2C19 PMs because of the increased probability of adverse effects ^15,43^. Appropriate support could be provided to patients at the beginning of the treatment to reduce the dropout rate and maximize the drug effect.

In *CYP2D6*, genotype data has lower ability to identify star alleles, severely compromising the assessment of pharmacogenetic effects. The lack of significant associations between metabolizer status and response is partly due to the limitations of genotype data to impute star alleles, and metabolizer status. Our study detected 16 star alleles of *CYP2D6*, which were classified as having no, decreased, or normal function. No structural variants could be detected, so increased function alleles (*xN) were not called, and fewer PMs/IMs were reported (such as undetected deletion *5). Approximately 7% of *CYP2D6* variants are structural variants, so the star allele calls, diplotype assignment and metabolic phenotype will be affected by missing structural variants ^22,44^. In contrast to a recent meta-analysis of clinical trials showing strong associations of CYP2D6-guided antidepressant treatment with improved patient outcomes ^45^, we found no association between CYP2D6 metabolizer status and treatment outcome in all samples or in the CYP2D6-antidepressant group. The low concordance rate of CYP2D6 metabolic phenotype with previous assessment in the GENDEP study ^17^ (79.9%) indicates a limited allele coverage in the genotype, leading to a higher proportion of NMs and reduced effect size of other metabolic phenotypes. Thus, our results should be interpreted with caution.

Activity score was also applied for the assignment of metabolic phenotype. Using clinical guidelines, each allele from *CYP2C19* and *CYP2D6* is assigned an activity value and the value is summed across the two alleles carried to give an activity score representing the individual’s metabolic activity ^34,35,40^. We found no effect of activity score on the outcomes of remission and percentage improvement but not the limitations of performing this across all drugs. These antidepressant results contrast to antipsychotic response, where higher CYP2C19 activity score was associated with lower symptom severity ^34^. The previous association of CYP2C19 PMs with remission outcome was not detected in the activity score analysis. This is likely because PMs have a low frequency and represent only the lower tail of the activity score distribution, so the effect is diluted when combining phenotype groups.

Our analyses included both European and East Asian ancestry populations. The frequencies of star alleles were clustered by ancestry. For example, European population had lower frequencies of *2, *3 in *CYP2C19* and *10 in *CYP2D6*, but higher frequencies of *CYP2C19* *17 and *CYP2D6* *4, than the East Asian population, leading to fewer PMs and IMs for CYP2C19 but higher proportions of CYP2C19 RMs+UMs and CYP2D6 PMs. These ancestry differences align with the CPIC guideline and other reports ^15,46^. When connecting the cytochrome enzyme status with antidepressant response, few studies have been performed in the East Asian population. A clinical trial of 100 depression patients from Taipei found CYP2D6 intermediate metabolizers had higher frequency of remitters and CYP2C19 poor metabolizers had higher serum levels of antidepressants ^47^. In some antipsychotics metabolized by specific cytochrome enzymes, the plasma concentrations of drugs are higher in East Asian populations than in European populations ^48^. In contrast, modelling has suggested that the metabolic contributions of CYP2C19 on escitalopram would be similar across European and Asian populations ^49^. As there is little evidence of differentiation by ancestry, current clinical guidelines provide the same antidepressant dosing recommendations across populations ^15^. Our subgroup meta-analyses between the European and East Asian studies found no difference in metabolic effect for each phenotype of both genes but the low sample size in East Asian studies (9% of all samples) implies much reduced power compared to the European studies.

Some study limitations should be considered. In addition to the incomplete assessment of star alleles from genotype data considered above, larger sample sizes are needed specifically in different ancestries and drug groups to evaluate drug-specific metabolic effect. Too few CYP2C19 PMs (2.1% in European, 3.1% in all participants) were present to show a statistically significant association after correcting for multiple testing. Similarly, CYP2C19 RMs+UMs (1.1%) and CYP2D6 PMs (< 0.1%) were rare in the East Asian population. Citalopram and escitalopram were the most prescribed drugs, accounting for 54% of all samples and 93% of the CYP2C19 antidepressant group, so the metabolic effect on treatment response was mainly determined by these two drugs. In addition, no data from clinical evaluations or the environment (e.g., dosage, concomitant drugs, smoking, diet) were analysed, and these factors could influence symptom improvement and cytochrome metabolic activity. Although no significant heterogeneity was detected in the meta-analyses, we should acknowledge the differences among cohorts in study design, patient selection, and response measures. No significant differences between IMs/RMs+UMs and NMs could be detected, and higher power is probably needed to effectively test between metabolizer groups. Side effects were also not available in our data, which are associated with metabolic phenotypes. We analysed only the final depression score, at the end of the study treatment, to determine remission and calculate the percentage improvement. Other studies have suggested using longitudinal measures throughout treatment period as repeated measures in a mixed linear model to improve the statistical power ^41^. While our study concentrates on *CYP2C19* and *CYP2D6*, there is potential for extending pharmacogenetic testing to other pharmacokinetic (*CYP2B6*) and pharmacodynamic genes (*SLC6A4, HTR2A*) to understand their impact on antidepressant efficacy and tolerability^15^. The combined effect of CYP2C19 and CYP2D6 on drugs metabolized by both genes (such as amitriptyline) could be explored for additional dosing recommendation. Finally, the imputed genotype showed a promising utility in detecting *CYP2C19* star alleles, but in *CYP2D6*, no RMs, UMs and fewer PMs/IMs were identified due to undetected rare and structural variants, increasing the false negative rate of CYP2D6 findings. Deeper imputation panels that detect structure variants, or further genetic studies using sequencing or modern targeted array would be necessary for a full assessment of CYP2D6 metabolizer status ^50^.

In conclusion, using imputed genotype data, our meta-analysis showed no significant association between cytochrome metabolic phenotypes with antidepressant response. Moderate evidence of an association with CYP2C19 poor metabolizers was indicated, which had higher rates of antidepressant remission. Metabolic phenotypes differed in frequency between European and East Asian populations but did not differ in their effect on treatment outcomes. Research studies with genotype are limited in their assessment of pharmacogenetic variation especially for CYP2D6. Fuller assessment of metabolizer status together with information on clinical factors and broader ancestry diversity could reduce the heterogeneity and improve power to evaluate the effect of metabolic phenotypes on antidepressant response.

## Data Availability

The data that support the findings of this study are available upon request.

## Acknowledgements

The collection of the sample from the Group for the Study of Resistant Depression (GSRD) Consortium was supported by an unrestricted grant from Lundbeck for the GSRD. Lundbeck had no further role in the study design and the collection, analysis, and interpretation of data.

The GENDEP (Genome Based Therapeutic Drugs for Depression) study was funded by a European Commission Framework 6 grant (EC Contract Ref. No. LSHB-CT-2003-503428). H. Lundbeck provided nortriptyline and escitalopram for the GENDEP study. GlaxoSmithKline and the UK National Institute for Health Research of the Department of Health contributed to the funding of the sample collection at the Institute of Psychiatry, London. GENDEP Illumina array genotyping was funded in part by a joint grant from the UK Medical Research Council and GlaxoSmithKline (Grant No. G0701420).

The GENPOD (GENetic and clinical Predictors Of treatment response in Depression) trial was funded by the UK Medical Research Council and supported by the UK Mental Health Research Network. The genotyping of GENPOD samples was supported by the Innovative Medicine Initiative Joint Undertaking under Grant No. 115008, of which resources are composed of European Union and the European Federation of Pharmaceutical Industries and Associations (EFPIA) in-kind contribution and financial contribution from the European Union’s Seventh Framework Programme (Grant No. FP7/2007-2013). EFPIA members Pfizer, GlaxoSmithKline, and F. Hoffmann-La Roche have contributed work and samples to the project presented here. The PFZ (Pfizer), GSK (GlaxoSmithKline), and GODS were supported by the Innovative Medicine Initiative Joint Undertaking (IMI-JU) under Grant No. 115008 of which resources are composed of European Union and EFPIA) in-kind contribution and financial contribution from the European Union’s Seventh Framework Programme (FP7/2007-2013). EFPIA members Pfizer, GlaxoSmithKline, and F. Hoffmann La-Roche have contributed work and samples to the project presented here.

The PGRN-AMPS (Pharmacogenomics Research Network Antidepressant Medication Pharmacogenomic Study) study data were obtained via the database of Genotypes and Phenotypes (dbGAP) (Accession No. phs000670.v1.p1). Funding support for the PGRN-AMPS study was provided by the National Institute of General Medical Sciences, National Institutes of Health, through the PGRN grant to Principal Investigators R. Weinshilboum and L. Wang (Grant No. U19 GM61388). D. Mrazek served as the Principal Investigator for the PGRN-AMPS study within the Mayo Clinic PGRN program. Genome-wide genotyping was performed at the RIKEN Center for Genomic Medicine, with funding provided by RIKEN. The datasets used for the analyses described in this manuscript were obtained from the dbGaP at http://www.ncbi.nlm.nih.gov/gap/.

Major funding for the Psychiatric Genetics Consortium (PGC) is from the U.S. National Institutes of Health (Grant Nos. U01 MH109528 and U01 MH109532).

Statistical analyses for the PGC were carried out on the NL Genetic Cluster Computer (http://www.geneticcluster.org) hosted by SURFsara.

For the purposes of open access, the author has applied a Creative Commons Attribution (CC BY) licence to any Accepted Author Manuscript version arising from this submission.

## Conflict of Interest

CML has served on the scientific advisory board for Myriad Neuroscience, and is a consultant for UCB. AS is or has been consultant/speaker for: Abbott, AbbVie, Angelini, AstraZeneca, Clinical Data, Boehringer, Bristol Myers Squibb, Eli Lilly, GlaxoSmithKline, InnovaPharma, Italfarmaco, Janssen, Lundbeck, Naurex, Pfizer, Polifarma, Sanofi, and Servier. AMM has received research support from the Sackler Trust and speakers fees from Janssen and Illumina.

MK has received grant funding from the Japanese Ministry of Health, Labor and Welfare, the Japan Society for the Promotion of Science, SENSHIN Medical Research Foundation, the Japan Research Foundation for Clinical Pharmacology and the Japanese Society of Clinical Neuropsychopharmacology and speaker’s honoraria from Sumitomo Pharma, Otsuka, Meiji-Seika Pharma, Eli Lilly, MSD K.K., Pfizer, Janssen Pharmaceutical, Shionogi, Mitsubishi Tanabe Pharma, Takeda Pharmaceutical, Lundbeck Viatris Inc, Eisai Co., Ltd. and Ono Pharmaceutical and participated in an advisory/review board for Otsuka, Sumitomo Pharma, Shionogi and Boehringer Ingelheim.

DS has received grant/research support from GlaxoSmithKline and Lundbeck; and served as a consultant or on advisory boards for AstraZeneca, Bristol Myers Squibb, Eli Lilly, Janssen, and Lundbeck.

CF was a speaker for Janssen.

NP is or has been consultant/speaker for: Takeda, Janssen and Lundbeck

All other authors report no biomedical financial interests or potential conflicts of interest.

## Data and code availability

The data that support the findings of this study are available upon request. Analysis code is available on the github: https://github.com/DanyangLi107/PGC_CYP2gene.

