## Supplementary for "Meta-analysis of CYP2C19 and CYP2D6 metabolic activity on antidepressant response from 13 clinical studies using genotype imputation"

**Li D, Pain O, Fabbri C et al.**

### Supplementary Materials

### Supplementary Author List

European Group for the Study of Resistant Depression (GSRD) Consortium

Siegfried Kasper,[1] Joseph Zohar,[2] Daniel Souery,[3] Stuart Montgomery,[4] Diego Albani,[5] Gianluigi Forloni,[5] Panagiotis Ferentinos,[6] Dan Rujescu,[7] Julien Mendlewicz.[8]

[1]Department of Psychiatry and Psychotherapy, Medical University Vienna, Austria;
[2]Department of Psychiatry, Sheba Medical Center, Tel Hashomer, and Sackler School of Medicine, Tel Aviv University, Israel;
[3]Laboratoire de Psychologie Médicale, Université Libre de Bruxelles and Psy Pluriel, Centre Européen de Psychologie Médicale, Brussels;
[4]Imperial College School of Medicine, London, UK;
[5]Laboratory of Biology of Neurodegenerative Disorders, Neuroscience Department, Istituto di Ricerche Farmacologiche Mario Negri IRCCS, Milan, Italy;
[6]Department of Psychiatry, Athens University Medical School, Athens, Greece;
[7]University Clinic for Psychiatry, Psychotherapy and Psychosomatic, Martin-Luther-University Halle-Wittenberg, Germany;
[8]Université Libre de Bruxelles.

**Major Depressive Disorder Working Group of the Psychiatric Genomics Consortium (2023)**

Mark J Adams 1 *
Fabian Streit 2 *
Swapnil Awasthi 3 *
Brett N Adey 4
Karmel W Choi 5, 6
V Kartik Chundru 7
Jonathan RI Coleman 4, 8
Jerome C Foo 2
Olga Giannakopoulou 9
Alisha S M Hall 2, 10
Jens Hjerling-Leffler 11
David M Howard 4
Christopher Hübel 4, 12, 13
Alex S F Kwong 1, 14
Bochao Danae Lin 15
Xiangrui Meng 9
Guiyan Ni 16
Oliver Pain 17
Gita A Pathak 18, 19
Eva C Schulte 20, 21, 22, 23
Jackson G Thorp 24
Alicia Walker 16
Shuyang Yao 25
Jian Zeng 16
Johan Zvrskovec 4, 8
Dag Aarsland 26
Ky'Era V Actkins 27
Mazda Adli 3, 28
Esben Agerbo 12, 29, 30
Mareike Aichholzer 31
Tracy M Air 32
Allison Aiello 33
Thomas D Als 30, 34, 35
Evelyn Andersson 36
Till F M Andlauer 37, 38
Volker Arolt 39
Helga Ask 40, 41
Sunita Badola 42
Clive Ballard 43
Karina Banasik 44
Nicholas J Bass 9
Aartjan T F Beekman 45
Sintia Belangero 46
Elisabeth B Binder 38, 47
Ottar Bjerkeset 48, 49
Gyda Bjornsdottir 50
Julia Boberg 36
Sigrid Børte 51, 52, 53
Emma Bränn 54
Alice Braun 55
Thorsten Brodersen 56
Søren Brunak 44
Mie T Bruun 57
Pichit Buspavanich 58, 59
Jonas Bybjerg-Grauholm 60, 61
Enda M Byrne 62
Archie Campbell 63, 64
Megan L. Campbell 65
Enrique Castelao 66
Jorge Cervilla 67, 68
Boris Chaumette 69
Chia-Yen Chen 70
Zhengming Chen 71, 72
Sven Cichon 73, 74, 75, 76
Lucía Colodro-Conde 24
Anne Corbett 43
Elizabeth C Corfield 40, 77
Baptiste Couvy-Duchesne 78
Nick Craddock 79, 80
Udo Dannlowski 39
Gail Davies 81
EJC de Geus 82
Ian J Deary 81
Franziska Degenhardt 76, 83
Abbas Dehghan 84, 85
J Raymond DePaulo 86
Michael Deuschle 87
Maria Didriksen 88
Khoa Manh Dinh 89
Nese Direk 90
Srdjan Djurovic 91, 92
Anna R Docherty 93, 94, 95
Katharina Domschke 96
Joseph Dowsett 88
Ole Kristian Drange 49, 97, 98, 99
Erin C Dunn 6, 100
Gudmundur Einarsson 50
Thalia C Eley 4
Samar S M Elsheikh 101
Jan Engelmann 102
Michael E Benros 60, 103, 104
Christian Erikstrup 89
Valentina Escott-Price 80
Chiara Fabbri 4, 105
Yu Fang 106
Sarah Finer 107
Josef Frank 2
Robert C Free 108
He Gao 109
Michael Gill 110
Maria Gilles 87
Fernando S Goes 86
Scott Douglas Gordon 24
Jakob Grove 30, 34, 35, 111
Daniel F Gudbjartsson 50, 112
Blanca Gutierrez 67, 68
Tim Hahn 39
Lynsey S Hall 80
Thomas F Hansen 44, 60, 113
Magnus Haraldsson 114
Catherina A Hartman 115
Alexandra Havdahl 40
Caroline Hayward 116
Stefanie Heilmann-Heimbach 76
Stefan Herms 74, 76
Ian B Hickie 117
Henrik Hjalgrim 118
Per Hoffmann 74, 76
Georg Homuth 119
Carsten Horn 120
Jouke-Jan Hottenga 82
David M Hougaard 60, 61
Iiris Hovatta 121
Qin Qin Huang 7
Floris Huider 82
Karen A Hunt 122
Marcus Ising 123
Erkki Isometsä 124
Rick Jansen 45
Yunxuan Jiang 125
Ian Jones 80
Lisa A Jones 126
Lina Jonsson 127
Robert Karlsson 25
Siegfried Kasper 128
Kenneth S Kendler 129
Ronald C Kessler 130
Stefan Kloiber 101, 123, 131, 132
James A Knowles 133
Nastassja Koen 65
Julia Kraft 55
Henry R Kranzler 134, 135
Kristi Krebs 136
Theodora Kunovac Kallak 137
Zoltán Kutalik 138, 139, 140
Elisa Lahtela 141
Margit Hørup Larsen 88
Eric J Lenze 142
Daniel F Levey 143, 144
Melissa Lewins 1
Glyn Lewis 9
Liming Li 145, 146
Kuang Lin 71
Penelope A Lind 24
Donald J MacIntyre 1, 147, 148
Dean F MacKinnon 86
Hermine HM Maes 149, 150
Wolfgang Maier 151
Victoria S Marshe 101, 152
Hamdi Mbarek 82
Peter McGuffin 4
Sarah E Medland 24
Susanne Meinert 39, 153
Susan Mikkelsen 89
Christina Mikkelsen 88, 154
Yuri Milaneschi 45
Iona Y Millwood 71, 72
Brittany L Mitchell 24
Esther Molina 67, 155
Francis M Mondimore 86
Preben Bo Mortensen 12, 29, 30
Benoit H Mulsant 101, 131
Joonas Naamanka 121

Jake M Najman 156
Matthias Nauck 157, 158
Igor Nenadi? 159
Kasper R Nielsen 160
Ilja M Nolte 161
Merete Nordentoft 60, 103, 104
Markus M Nöthen 76
Mette Nyegaard 30, 162, 163, 164
Michael C O'Donovan 80
Asmundur Oddsson 50
Catherine M Olsen 165, 166
Hogni Oskarsson 167
Sisse Rye Ostrowski 88, 168
Vanessa K Ota 46
Michael J Owen 80
Richard Packer 169
Teemu Palviainen 141
Pedro M Pan 170
Carlos N Pato 171
Michele T Pato 171
Nancy L Pedersen 25
Ole Birger Pedersen 172
Roseann E Peterson 129, 173
Wouter J Peyrot 45
James B Potash 86
Martin Preisig 66
Jorge A Quiroz 174
Charles F Reynolds III 175
John P Rice 142
Giovanni A Salum 176
Robert A Schoevers 177, 178
Andrew Schork 30, 179, 180
Thomas G Schulze 2, 21, 86, 181, 182
Tabea S Send 87
Jianxin Shi 183
Engilbert Sigurdsson 114
Kritika Singh 27
Grant C B Sinnamon 184
Lea Sirignano 2
Olav B Smeland 185, 186
Daniel J Smith 187
Erik Sørensen 88
Sundararajan Srinivasan 188
Hreinn Stefansson 50
Kari Stefansson 50, 189
Dan J. Stein 190
Frederike Stein 191
André Tadic 102, 192
Henning Teismann 193
Alexander Teumer 194
Anita Thapar 80, 195
Patrick F Sullivan 25, 253
Martin Tesli 40
Thorgeir E Thorgeirsson 50
Henning Tiemeier 254, 255
Pippa A Thomson 64
Lise Wegner Thørner 88
Apostolia Topaloudi 196
Ioanna Tzoulaki 84, 85, 197
Monica Uddin 198
André G Uitterlinden 199
Henrik Ullum 88, 200, 201
Daniel Umbricht 202
Robert J Ursano 203
Sandra Van der Auwera 204
David A van Heel 122
Albert M van Hemert 205
Abirami Veluchamy 188
Alexander Viktorin 25
Henry Völzke 194
Agaz Wani 198
G Bragi Walters 50
Robin G Walters 71, 72
Sylvia Wassertheil-Smoller 206
Myrna M Weissman 207, 208
Jürgen Wellmann 193
David C Whiteman 165
Derek Wildman 198
Gonneke Willemsen 82
Alexander T Williams 169
Bendik S Winsvold 51, 52, 209
Stephanie H Witt 2
Ying Xiong 25
Lea Zillich 2
John-Anker Zwart 51, 52, 53
23andMe Research Team 125
Estonian Biobank Research Team 136
HUNT All-In Psychiatry 210
China Kadoorie Biobank Collaborative Group 211
Genes & Health Research Team 212
Ole A Andreassen 185, 186, 213
Bernhard T Baune 214, 215, 216
Klaus Berger 193
Dorret I Boomsma 82
Anders D Børglum 30, 34, 35
Gerome Breen 4, 8
Na Cai 217, 218, 219
Hilary Coon 94
William E Copeland 220
Byron Creese 43
Lea K Davis 27
Eske M Derks 24
Enrico Domenici 221
Paul Elliott 84, 85, 197, 222
Andreas J Forstner 73, 76
Micha Gawlik 223
Joel Gelernter 19, 143, 224
Hans J Grabe 204
Steven P Hamilton 225
Kristian Hveem 226, 227, 228
Catherine John 169, 229
Jaakko Kaprio 141
Tilo Kircher 159
Marie-Odile Krebs 230
Karoline Kuchenbaecker 9, 71
Mikael Landén 25, 127
Kelli Lehto 136
Douglas F Levinson 231
Qingqin S Li 232
Klaus Lieb 102
Yi Lu 25
Susanne Lucae 123
Jurjen J Luykx 15, 233
Patrik K Magnusson 25
Nicholas G Martin 24
Hilary C Martin 7
Andrew McQuillin 9
Christel M Middeldorp 62, 234
Lili Milani 136
Ole Mors 30, 235
Daniel J Müller 101, 131, 132, 236
Bertram Müller-Myhsok 38, 237, 238
Albertine J Oldehinkel 115
Sara A Paciga 239
Colin NA Palmer 188
Peristera Paschou 196
Brenda WJH Penninx 45
Roy H Perlis 5, 6, 240
Giorgio Pistis 66
Renato Polimanti 18, 19
David J Porteous 64
Danielle Posthuma 241, 242
Ted Reichborn-Kjennerud 40
Andreas Reif 31
Frances Rice 80, 243
Roland Ricken 3
Marcella Rietschel 2
Margarita Rivera 67, 244
Christian Rück 245
Catherine Schaefer 246
Srijan Sen 106, 247
Alessandro Serretti 105
Alkistis Skalkidou 137
Jordan W Smoller 5, 248, 249
Frederike Stein 191
Murray B Stein 250, 251, 252

Nicholas J Timpson 14
Rudolf Uher 256
Jens R Wendland 42
Thomas Werge 60, 179, 201, 257, 258
Naomi R Wray 16, 259 **
Stephan Ripke 3, 248 **
Cathryn M Lewis 4, 260 **
Andrew M McIntosh 1, 261 **

* Joint Lead Authors
** Joint Last Authors

1, Division of Psychiatry, University of Edinburgh, Edinburgh, UK
2, Department of Genetic Epidemiology in Psychiatry, Central Institute of Mental Health, Medical Faculty Mannheim, Heidelberg University, Mannheim, BW, DE
3, Department of Psychiatry and Psychotherapy, Charité – Universitätsmedizin Berlin, Berlin, BE, DE
4, Social, Genetic and Developmental Psychiatry Centre, King's College London, London, UK
5, Department of Psychiatry, Massachusetts General Hospital, Boston, MA, US
6, Department of Psychiatry, Harvard Medical School, Boston, MA, US
7, Human Genetics, Wellcome Sanger Institute, Hinxton, UK
8, NIHR Maudsley Biomedical Research Centre, King's College London, London, UK
9, Division of Psychiatry, University College London, London, UK
10, Department of Clinical Medicine, Aarhus University, Aarhus, DK
11, Department of Medical Biochemistry and Biophysics, Karolinska Institutet, Stockholm, SE
12, National Centre for Register-based Research, Aarhus University, Aarhus, DK
13, Department of Pediatric Neurology, Charité – Universitätsmedizin Berlin, Berlin, BE, DE
14, MRC Integrative Epidemiology Unit, University of Bristol, Bristol, UK
15, Department of Psychiatry and Neuropsychology, School for Mental Health and Neuroscience, Maastricht University Medical Centre, Maastricht, NL
16, Institute for Molecular Bioscience, University of Queensland, Brisbane, QLD, AU
17, Maurice Wohl Clinical Neuroscience Institute, Department of Basic and Clinical Neuroscience, King's College London, London, UK
18, Veterans Affairs Connecticut Healthcare System, West Haven, CT, US
19, Department of Psychiatry, Yale University School of Medicine, New Haven, CT, US
20, Department of Psychiatry, University of Munich, Munich, BY, DE
21, Institute of Psychiatric Phenomics and Genomics, University of Munich, Munich, BY, DE
22, Department of Psychiatry and Psychotherapy, University Hospital Bonn, Medical Faculty, University of Bonn, Bonn, DE
23, Institute of Human Genetics, University Hospital Bonn, Medical Faculty, University of Bonn, Bonn, DE
24, Mental Health and Neuroscience, QIMR Berghofer Medical Research Institute, Brisbane, QLD, AU
25, Department of Medical Epidemiology and Biostatistics, Karolinska Institutet, Stockholm, SE
26, Old Age Psychiatry, King's College London, London, UK
27, Department of Medicine, Division of Genetic Medicine, Vanderbilt University Medical Center, Nashville, TN, US
28, Department of Psychiatry and Psychotherapy, Fliedner Klinik Berlin, Berlin, BE, DE
29, Centre for Integrated Register-based Research, Aarhus University, Aarhus, DK
30, iPSYCH, The Lundbeck Foundation Initiative for Integrative Psychiatric Research, Aarhus, DK
31, Department of Psychiatry, Psychosomatic Medicine and Psychotherapy, Goethe University Frankfurt - University Hospital, Frankfurt am Main, DE
32, Discipline of Psychiatry, University of Adelaide, Adelaide, SA, AU
33, Department of Epidemiology, Columbia University Mailman School of Public Health, New York, NY, US
34, Department of Biomedicine and Centre for Integrative Sequencing, iSEQ, Aarhus University, Aarhus, DK
35, Center for Genomics and Personalized Medicine, Aarhus University, Aarhus, DK
36, Department of Clinical Neuroscience, Karolinska Institutet,, SE
37, Department of Neurology, Klinikum rechts der Isar, Technical University of Munich, Munich, BY, DE
38, Department of Translational Research in Psychiatry, Max Planck Institute of Psychiatry, Munich, BY, DE
39, Institute for Translational Psychiatry, University of Münster, Münster, NRW, DE
40, Department of Mental Disorders, Norwegian Institute of Public Health, Oslo, NO
41, PROMENTA Research Center, Department of Psychology, University of Oslo, Oslo, NO
42, Research and Development, Takeda Pharmaceutical Company Limited, Cambridge, MA, US
43, Faculty of Health and Life Sciences, University of Exeter, Exeter, UK
44, Novo Nordisk Center for Protein Research, Department of Health Sciences, University of Copenhagen, Copenhagen, DK
45, Department of Psychiatry, Amsterdam Public Health and Amsterdam Neuroscience, Amsterdam UMC, Vrije Universiteit Amsterdam, Amsterdam, NL
46, Morphology and Genetics, Universidade Federal de Sao Paulo, Sao Paulo, SP, BR
47, Department of Psychiatry and Behavioral Sciences, Emory University School of Medicine, Atlanta, GA, US
48, Faculty of Nursing and Health Sciences, NORD University, Levanger, NO
49, Department of Mental Health, Faculty of Medicine and Health Sciences, Norwegian University of Science and Technology (NTNU), Trondheim, TRD, NO

50, deCODE Genetics / Amgen, Reykjavik, IS
51, K. G. Jebsen Center for Genetic Epidemiology, Department of Public Health and Nursing, Faculty of Medicine and Health Sciences, Norwegian University of Science and Technology (NTNU), Trondheim, TRD, NO
52, Department of Research and Innovation, Division of Clinical Neuroscience, Oslo University Hospital, Oslo, NO
53, Institute of Clinical Medicine, Faculty of Medicine, University of Oslo, Oslo, NO
54, Institute of Environmental Medicine, Unit of Integrative Epidemiology, Karolinska Institutet, Stockholm, SE
55, Department of Psychiatry and Psychotherapy, Charité – Universitätsmedizin Berlin, Berlin, DE
56, Department of Clinical Immunology, Roskilde University/Næstved Hospital, Roskilde, DK
57, Department of Clinical Immunology, Odense University Hospital, Odense, DK
58, Department of Psychiatry, Psychotherapy and Psychosomatics, Brandenburg Medical School Theodor Fontane, Neuruppin, BB, DE
59, Department of Psychiatry and Psychotherapy, Gender Research in Medicine, Institute of Sexology and Sexual Medicine, Charité – Universitätsmedizin Berlin, Berlin, BE, DE
60, iPSYCH, The Lundbeck Foundation Initiative for Integrative Psychiatric Research, Copenhagen, DK
61, Center for Neonatal Screening, Department for Congenital Disorders, Statens Serum Institut, Copenhagen, DK
62, Child Health Research Centre, University of Queensland, Brisbane, QLD, AU
63, Centre for Medical Informatics, Usher Institute, University of Edinburgh, Edinburgh, UK
64, Centre for Genomic & Experimental Medicine, Institute for Genetics and Cancer, University of Edinburgh, Edinburgh, UK
65, Department of Psychiatry and Mental Health, University of Cape Town, Cape Town, SA
66, Department of Psychiatry, Lausanne University Hospital and University of Lausanne, Prilly, VD, CH
67, Instituto de Investigación Biosanitaria ibs.GRANADA, Granada, ES
68, Department of Psychiatry, Faculty of Medicine and Institute of Neurosciences, Biomedical Research Centre (CIBM), University of Granada, Granada, ES
69, Université de Paris Cité, INSERM U1266, Institute of Psychiatry and Neuroscience of Paris, GHU Paris Psychiatry and Neuroscience, Paris, FR
70, Translational Biology, Biogen, Cambridge, MA, US
71, Nuffield Department of Population Health, University of Oxford, Oxford, UK
72, MRC Population Health Research Unit, University of Oxford, Oxford, UK
73, Institute of Neuroscience and Medicine (INM-1), Research Center Juelich, Juelich, DE
74, Human Genomics Research Group, Department of Biomedicine, University of Basel, Basel, CH
75, Institute of Medical Genetics and Pathology, University Hospital Basel, University of Basel, Basel, CH
76, Institute of Human Genetics, University of Bonn, School of Medicine & University Hospital Bonn, Bonn, DE
77, Nic Waals Institute, Lovisenberg Diakonale Hospital, Oslo, NO
78, Centre for Advanced Imaging, University of Queensland, Saint Lucia, QLD, AU
79, Psychological Medicine, Cardiff University, Cardiff, WLS, UK
80, Centre for Neuropsychiatric Genetics and Genomics, Cardiff University, Cardiff, WLS, UK
81, The Lothian Birth Cohorts, University of Edinburgh, Edinburgh, UK
82, Department of Biological Psychology & Amsterdam Public Health Research Institute, Vrije Universiteit Amsterdam, Amsterdam, NL
83, Department of Child and Adolescent Psychiatry, Psychosomatics and Psychotherapy, University Hospital Essen, Unversity of Duisburg-Essen, Duisburg, DE
84, MRC Centre for Environment and Health, School of Public Health, Imperial College London, London, UK
85, Imperial College Dementia Research Institute, Imperial College London, London, UK
86, Department of Psychiatry and Behavioral Sciences, Johns Hopkins University School of Medicine, Baltimore, MD, US
87, Department of Psychiatry and Psychotherapy, Research Group Stress Related Disorders, Central Institute of Mental Health, Medical Faculty Mannheim, Heidelberg University, Mannheim, BW, DE
88, Department of Clinical Immunology, Copenhagen University Hospital, Rigshospitalet, Copenhagen, CPH, DK
89, Department of Clinical Immunology, Aarhus University Hospital, Aarhus, DK
90, Department of Psychiatry, Istanbul University, Istanbul, TR
91, Department of Medical Genetics, Oslo University Hospital, Oslo, OSL, NO
92, NORMENT, Department of Clinical Science, University of Bergen, Bergen, NO
93, Virginia Institute for Psychiatric & Behavioral Genetics, Virginia Commonwealth University, Richmond, VA, US
94, Psychiatry Department / Huntsman Mental Health Institute, University of Utah School of Medicine, Salt Lake City, UT, US
95, Center for Genomic Research, University of Utah School of Medicine, Salt Lake City, UT, US

96, Department of Psychiatry and Psychotherapy, Medical Center, University of Freiburg, Faculty of Medicine, University of Freiburg, Freiburg, DE
97, Division of Mental Health Care, St. Olavs Hospital, Trondheim University Hospital, Trondheim, TRD, NO
98, Department of Psychiatry, Sørlandet Hospital, Kristiansand, AG, NO
99, University of Oslo, NORMENT Centre, Institute of Clinical Medicine, Oslo, OSL, NO
100, Center for Genomic Medicine, Massachusetts General Hospital, Boston, MA, US
101, Centre for Addiction and Mental Health, Toronto, ON, CA
102, Department of Psychiatry and Psychotherapy, University Medical Center of the Johannes Gutenberg University Mainz, Mainz, DE
103, Mental Health Center Copenhagen, Mental Health Services Capital Region of Denmark, Copenhagen, DK
104, Faculty of Health Science, Department of Clinical Medicine, University of Copenhagen, Copenhagen, DK
105, Department of Biomedical and Neuromotor Sciences, University of Bologna, Bologna, IT
106, Michigan Neuroscience Institute, University of Michigan, Ann Arbor, MI, US
107, Wolfson Institute of Population Health, Queen Mary University of London, London, UK
108, School of Computing and Mathematical Sciences, University of Leicester, Leicester, UK
109, Department of Epidemiology and Biostatistics, Imperial College London, London, UK
110, Discipline of Psychiatry, School of Medicine, Trinity College Dublin, Dublin, IE
111, Bioinformatics Research Centre, Aarhus University, Aarhus, DK
112, School of Engineering, University of Iceland, Reykjavik, IS
113, Danish Headache Centre, Department of Neurology, Rigshospitalet, Glostrup, DK
114, Faculty of Medicine, Department of Psychiatry, University of Iceland, Reykjavik, IS
115, Department of Psychiatry, University of Groningen, University Medical Center Groningen, Groningen, NL
116, MRC Human Genetics Unit, Institute for Genetics and Cancer, University of Edinburgh, Edinburgh, UK
117, Brain and Mind Centre, University of Sydney, Sydney, NSW, AU
118, Department of Epidemiology Research, Statens Serum Institut, Copenhagen, DK
119, Interfaculty Institute for Genetics and Functional Genomics, Department of Functional Genomics, University Medicine Greifswald, Greifswald, MV, DE
120, Roche Pharmaceutical Research and Early Development, Pharmaceutical Sciences, Roche Innovation Center Basel, F. Hoffmann-La Roche Ltd, Basel, CH
121, SleepWell Research Program and Department of Psychology and Logopedics, University of Helsinki, Helsinki, FI
122, Blizard Institute, Barts and the London School of Medicine and Dentistry, Queen Mary University of London, London, UK
123, Max Planck Institute of Psychiatry, Munich, BY, DE
124, Department of Psychiatry, University of Helsinki, Helsinki, FI
125, 23andMe Research Team, 23andMe, Inc., Sunnyvale, CA, US
126, Department of Psychological Medicine, University of Worcester, Worcester, UK
127, Institution of Neuroscience and Physiology, University of Gothenburg, Gothenburg, SE
128, Department of Psychiatry and Psychotherapy, Medical University of Vienna, Vienna, AT
129, Department of Psychiatry, Virginia Commonwealth University, Richmond, VA, US
130, Health Care Policy, Harvard Medical School, Boston, MA, US
131, Department of Psychiatry, University of Toronto, Toronto, ON, CA
132, Department of Pharmacology & Toxicology, University of Toronto, Toronto, ON, CA
133, Department of Genetics, Rutgers University, Piscataway, NJ, US
134, Department of Psychiatry, Perelman School of Medicine, University of Pennsylvania, Philadelphia, PA, US
135, Mental Illness Research, Education and Clinical Center, Crescenz VA Medical Center, Philadelphia, PA, US
136, Estonian Genome Centre, Institute of Genomics, University of Tartu, Tartu, EE
137, Department of Women's and Children's Health, Uppsala University, Uppsala, SE
138, Department of Epidemiology and Health Systems, Center for Primary Care and Public Health, Lausanne, VD, CH
139, Swiss Institute of Bioinformatics, Lausanne, VD, CH
140, Department of Computational Biology, University of Lausanne, Lausanne, VD, CH
141, Institute for Molecular Medicine Finland - FIMM, University of Helsinki, Helsinki, FI
142, Department of Psychiatry, Washington University School of Medicine in St. Louis, St. Louis, MO, US
143, Psychiatry, Veterans Affairs Connecticut Healthcare System, West Haven, CT, US
144, Department of Psychiatry, Yale University, New Haven, CT, US
145, Department of Epidemiology and Biostatistics, School of Public Health, Peking University, Beijing, CN
146, Peking University Center for Public Health and Epidemic Preparedness & Response, Peking University, Beijing, CN
147, Mental Health, NHS 24, Glasgow, UK

148, Royal Edinburgh Hospital, NHS Lothian, Edinburgh, UK
149, Department of Human and Molecular Genetics, Virginia Commonwealth University, Richmond, VA, USA
150, Virginia Institute for Psychiatric and Behavioral Genetics, Virginia Commonwealth University, Richmond, VA, USA
151, Department of Psychiatry and Psychotherapy, University of Bonn, Bonn, DE
152, Center for Translational and Computational Neuroimmunology, Columbia University Medical Center, New York, NY, US
153, Institute for Translational Neuroscience, University of Münster, Münster, NRW, DE
154, Novo Nordisk Foundation Center for Basic Metabolic Research, Faculty of Health Science, Copenhagen University, Copenhagen, DK
155, Department of Nursing, Faculty of Health Sciences and Institute of Neurosciences, Biomedical Research Centre (CIBM), University of Granada, Granada, ES
156, School of Public Health, University of Queensland, Brisbane, QLD, AU
157, DZHK (German Centre for Cardiovascular Research), Partner Site Greifswald, Greifswald, MV, DE
158, Institute of Clinical Chemistry and Laboratory Medicine, University Medicine Greifswald, Greifswald, MV, DE
159, Department of Psychiatry, University of Marburg, Marburg, DE
160, Department of Clinical Immunology, Aalborg University Hospital, Aalborg, DK
161, Department of Epidemiology, University of Groningen, University Medical Center Groningen, Groningen, NL
162, Department of Health, Science and Technology, Aalborg University, Aalborg, DK
163, Centre for Integrative Sequencing, iSEQ, Aarhus University, Aarhus, DK
164, Department of Biomedicine-Human Genetics, Aarhus University, Aarhus, DK
165, Population Health, QIMR Berghofer Medical Research Institute, Brisbane, QLD, AU
166, The Fraser Institute, Faculty of Medicine, University of Queensland, Brisbane, QLD, AU
167, Humus, Reykjavik, IS
168, Department of Clinical Medicine, University of Copenhagen, Copenhagen, CPH, DK
169, Department of Population Health Sciences, University of Leicester, Leicester, UK
170, Department of Psychiatry, Universidade Federal de Sao Paulo, Sao Paulo, SP, BR
171, Department of Psychiatry, Rutgers University, Piscataway, NJ, US
172, Department of Clinical Immunology, Zealand University Hospital, Køge, DK
173, Department of Psychiatry and Behavioral Sciences, SUNY Downstate Health Sciences University, Brooklyn, NY, US
174, NMD Pharma, Lexington, MA, US
175, Psychiatry, University of Pittsburgh Medical Centre, Pittsburgh, PA, US
176, Psychiatry, Universidade Federal do Rio Grande do Sul, Porto Alegre, BR
177, Department of Psychiatry, University Medical Center Groningen, Groningen, NL
178, Research School of Behavioural and Cognitive Neurosciences (BCN), University of Groningen, Groningen, NL
179, Institute of Biological Psychiatry, Mental Health Center Sct. Hans, Mental Health Services Capital Region of Denmark, Copenhagen, DK
180, Neurogenomics Division, The Translational Genomics Research Institute (TGEN), Phoenix, AZ, US
181, Human Genetics Branch, NIMH Division of Intramural Research Programs, Bethesda, MD, US
182, Department of Psychiatry and Psychotherapy, University Medical Center Göttingen, Goettingen, NI, DE
183, Division of Cancer Epidemiology and Genetics, National Cancer Institute, Bethesda, MD, US
184, School of Medicine and Dentistry, James Cook University, Townsville, QLD, AU
185, Division of Mental Health and Addiction, Oslo University Hospital, Oslo, OSL, NO
186, NORMENT, Institute of Clinical Medicine, University of Oslo, Oslo, OSL, NO
187, Institute of Health and Wellbeing, University of Glasgow, Glasgow, UK
188, Division of Population Health and Genomics, Ninewells Hospital and School of Medicine, University of Dundee, Dundee, UK
189, Faculty of Medicine, University of Iceland, Reykjavik, IS
190, SAMRC Unit on Risk & Resilience in Mental Disorders, Department of Psychiatry and Mental Health, University of Cape Town, Cape Town, SA
191, Department of Psychiatry and Psychotherapy, University of Marburg, Marburg, HE, DE
192, Department of Psychiatry, Psychotherapy and Psychosomatics, Dr. Fontheim Mentale Gesundheit, Liebenburg, DE
193, Institute of Epidemiology and Social Medicine, University of Münster, Münster, NRW, DE
194, Institute for Community Medicine, University Medicine Greifswald, Greifswald, MV, DE
195, Wolfson Centre for Young People's Mental Health, Division of Psychological Medicine and Clinical Neurosciences, Cardiff University, Cardiff, WLS, UK
196, Department of Biological Sciences, Purdue University, West Lafayette, IN, US
197, Imperial College BHF Centre for Research Excellence, Imperial College London, London, UK

198, Genomics Program, University of South Florida College of Public Health, Tampa, FL, US
199, Department of Internal Medicine, Erasmus University Medical Center Rotterdam, Rotterdam, NL
200, Management Section, Statens Serum Institut, Copenhagen, DK
201, Department of Clinical Medicine, University of Copenhagen, Copenhagen, DK
202, Xperimed LLC, Basel, CH
203, Psychiatry, USUHS, Bethesda, US
204, Department of Psychiatry and Psychotherapy, University Medicine Greifswald, Greifswald, MV, DE
205, Department of Psychiatry, Leiden University Medical Center, Leiden, NL
206, Department of Epidemiology and Population Health, Albert Einstein College of Medicine, Bronx, NY, US
207, Department of Psychiatry, Columbia University College of Physicians and Surgeons, New York, NY, US
208, Division of Epidemiology, New York State Psychiatric Institute, New York, NY, US
209, Department of Neurology, Oslo University Hospital, Oslo, NO
210, HUNT All-In Psychiatry
211, China Kadoorie Biobank Collaborative Group
212, Genes & Health Research Team
213, KG Jebsen Centre for Neurodevelopmental Research, University of Oslo, Oslo, OSL, NO
214, Department of Psychiatry, University of Melbourne, Melbourne, VIC, AU
215, Florey Institute of Neuroscience and Mental Health, University of Melbourne, Melbourne, VIC, AU
216, Department of Psychiatry, University of Münster, Münster, NRW, DE
217, Computational Health Centre, Helmholtz Zentrum München, Neuherberg, DE
218, School of Medicine, Technical University of Munich, Munich, BY, DE
219, Helmholtz Pioneer Campus, Helmholtz Zentrum München, Neuherberg, DE
220, Department of Psychiatry, University of Vermont, Burlington, VT, US
221, Department of Cellular, Computational and Integrative Biology, Università degli Studi di Trento, Trento, IT
222, Imperial College Biomedical Research Centre, Imperial College London, London, UK
223, Department of Psychiatry, Psychosomatics and Psychotherapy, Julius-Maximilians-Universität Würzburg, Würzburg, DE
224, Department of Genetics, Department of Neuroscience, Yale University School of Medicine, New Haven, CT, US
225, Psychiatry, Kaiser Permanente Northern California, San Francisco, CA, US
226, K. G. Jebsen Center for Genetic Epidemiology, Department of Public Health and Nursing, Faculty of Medicine and Health Sciences, Norwegian University of Science and Technology (NTNU), Trondheim, NO
227, HUNT Research Center, Department of Public Health and Nursing, Faculty of Medicine and Health Sciences, Norwegian University of Science and Technology (NTNU), Trondheim, NO
228, Department of Research, Innovation and Education, St. Olavs Hospital, Trondheim University Hospital, Trondheim, NO
229, NIHR Leicester Biomedical Research Centre, Glenfield Hospital, Leicester, UK
230, Pathophysiology of Psychiatric Diseases, INSERM, Univ Paris Cité, GHU Paris, Paris, FR
231, Department of Psychiatry & Behavioral Sciences, Stanford University, Stanford, CA, US
232, Neuroscience Therapeutic Area, Janssen Research and Development, LLC, Titusville, NJ, US
233, Second Opinion Outpatient Clinic, GGNet Mental Health, Warnsveld, NL
234, Child and Youth Mental Health Service, Children's Health Queensland Hospital and Health Service, Brisbane, QLD, AU
235, Psychosis Research Unit, Aarhus University Hospital-Psychiatry, Aarhus, DK
236, Department of Psychiatry, Psychosomatics and Psychotherapy, University Hospital of Würzburg, Würzburg, DE
237, Munich Cluster for Systems Neurology (SyNergy), Munich, BY, DE
238, University of Liverpool, Liverpool, UK
239, Human Genetics and Computational Biomedicine, Pfizer Global Research and Development, Groton, CT, US
240, Centre for Quantitative Health, Massachusetts General Hospital, Boston, MA, US
241, Child and Adolescent Psychiatry, Amsterdam UMC, Vrije Universiteit Amsterdam, Amsterdam, NL
242, Complex Trait Genetics, Vrije Universiteit Amsterdam, Amsterdam, NL
243, Wolfson Centre for Young People's Mental Health, Division of Psychological Medicine and Clinical Neurosciences, Cardiff University, Cardiff, UK
244, Department of Biochemistry and Molecular Biology II, Faculty of Pharmacy and Institute of Neurosciences, Biomedical Research Centre (CIBM), University of Granada, Granada, ES
245, Department of Clinical Neuroscience, Karolinska Institutet, Stockholm, SE
246, Division of Research, Kaiser Permanente Northern California, Oakland, CA, US
247, Department of Psychiatry, University of Michigan, Ann Arbor, MI, US

248, Stanley Center for Psychiatric Research, Broad Institute of MIT and Harvard, Cambridge, MA, US
249, Psychiatric and Neurodevelopmental Genetics Unit, Massachusetts General Hospital, Boston, MA, US
250, Psychiatry, UCSD School of Medicine, La Jolla, CA, US
251, Public Health, UCSD School of Public Health, La Jolla, CA, US
252, Psychiatry, Veterans Affairs San Diego Healthcare System, San Diego, CA, US
253, Departments of Genetics and Psychiatry, University of North Carolina at Chapel Hill, Chapel Hill, NC, US
254, Child and Adolescent Psychiatry, Erasmus University Medical Center Rotterdam, Rotterdam, NL
255, Social and Behavioral Science, Harvard T.H. Chan School of Public Health, Boston, MA, US
256, Psychiatry, Dalhousie University, Halifax, NS, CA
257, Institute of Biological Psychiatry, Mental Health Center Sct. Hans, Copenhagen University Hospital, Mental Health Services, Copenhagen, DK
258, GLOBE Institute, Lundbeck Foundation Centre for Geogenetics, University of Copenhagen, Copenhagen, DK
259, Queensland Brain Institute, University of Queensland, Brisbane, QLD, AU
260, Department of Medical & Molecular Genetics, King's College London, London, UK
261, Institute for Genomics and Cancer, University of Edinburgh, Edinburgh, UK

**Supplementary Figures**

Supplementary Figure 1. Distribution of CYP2C19 and CYP2D6 antidepressants in each cohort

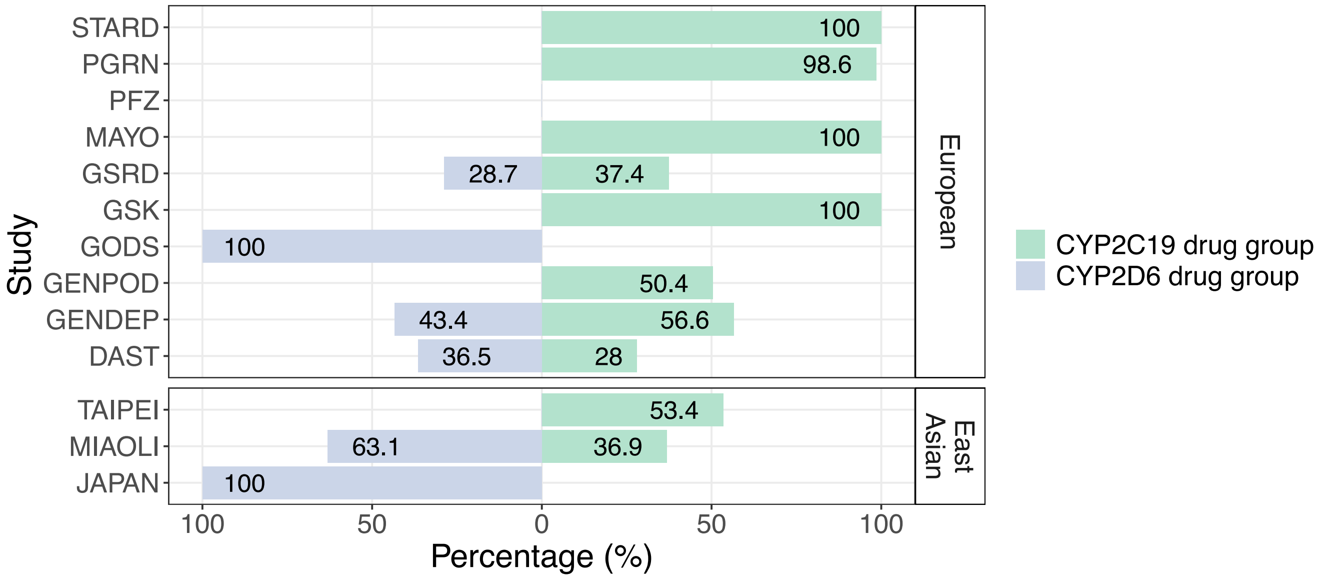

Supplementary Figure 2. Frequency of CYP2C19 and CYP2D6 star alleles

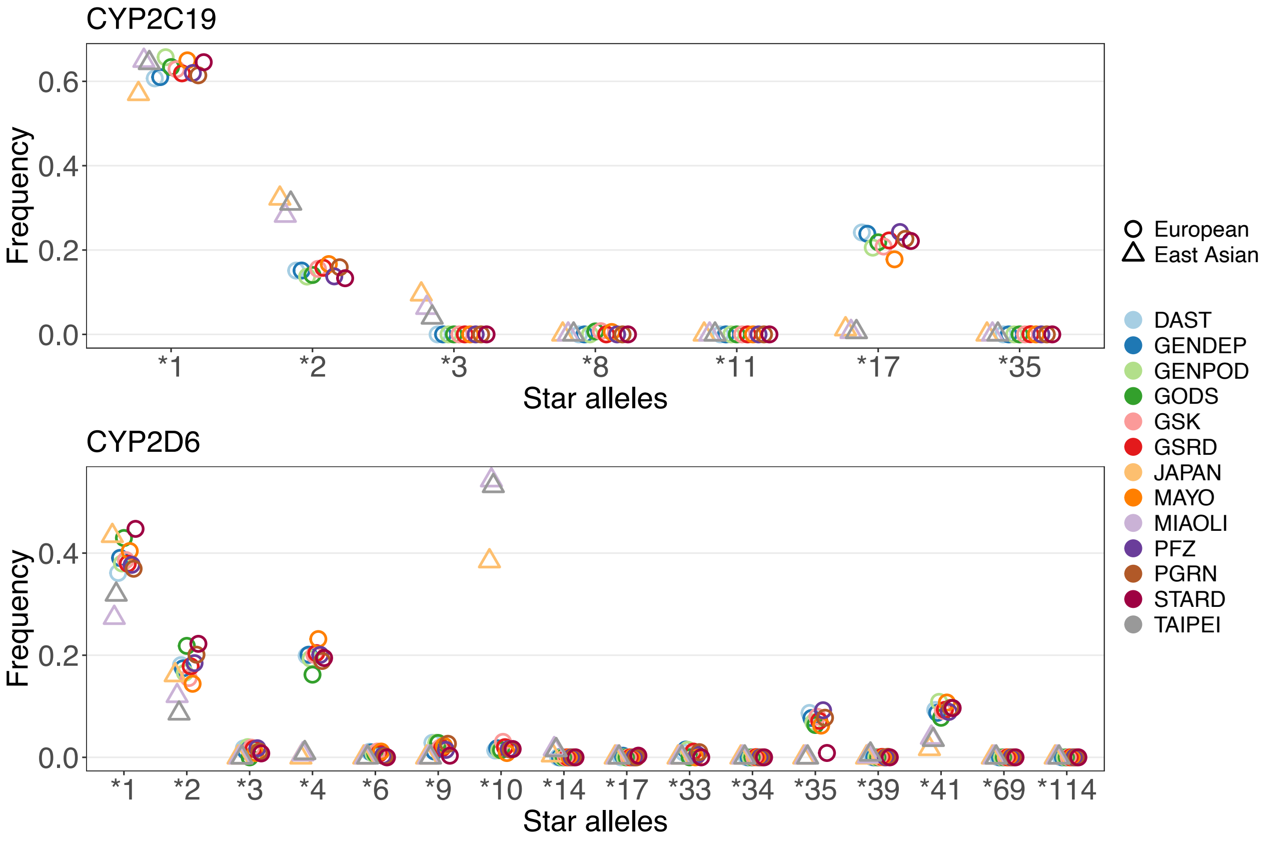

Supplementary Figure 3. Meta-analyses of CYP2C19 poor, intermediate, and ultrarapid metabolizers in all samples

### CYP2C19 poor metabolizers

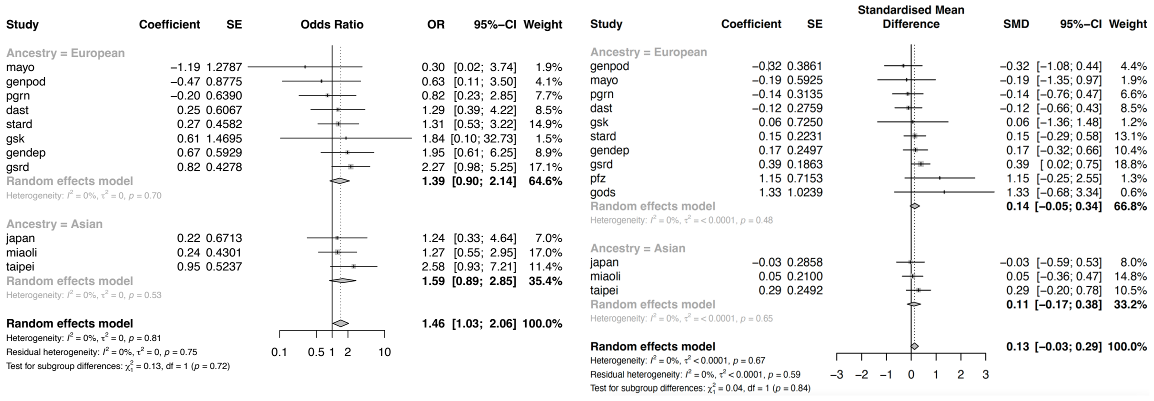

### CYP2C19 intermediate metabolizers

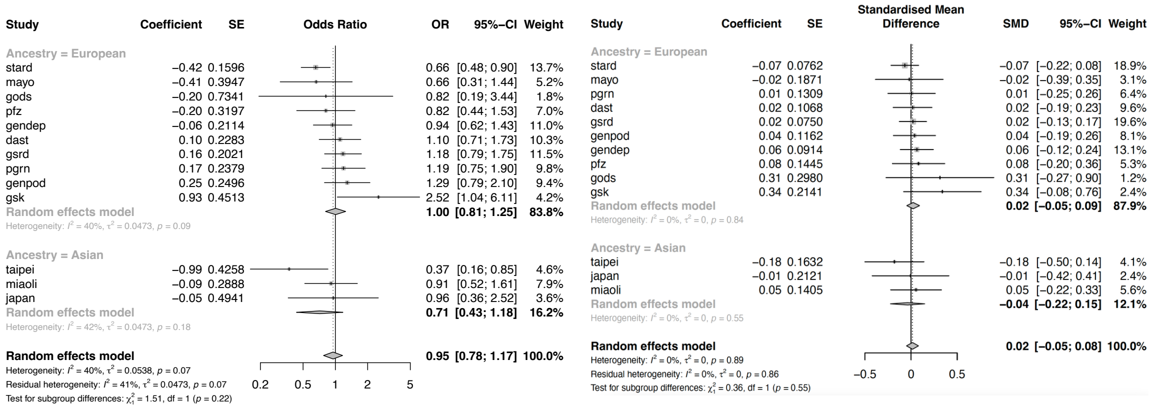

### CYP2C19 ultrarapid metabolizers

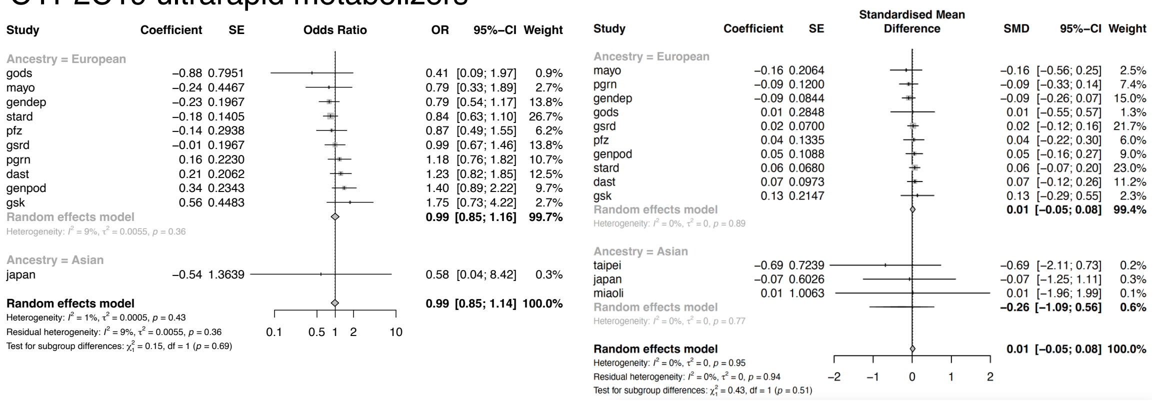

SE: standard deviation, CI: confidence interval

Supplementary Figure 4. Meta-analyses of CYP2D6 poor and intermediate metabolizers in all samples

### CYP2D6 poor metabolizers

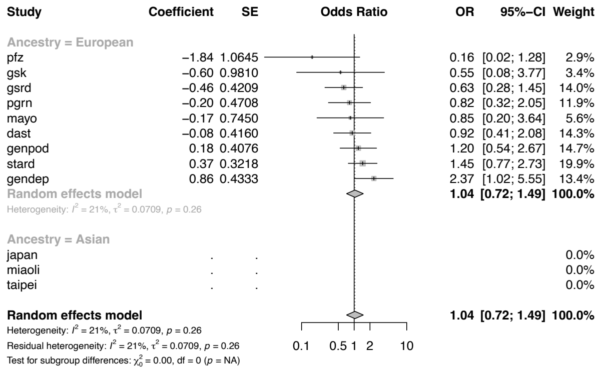

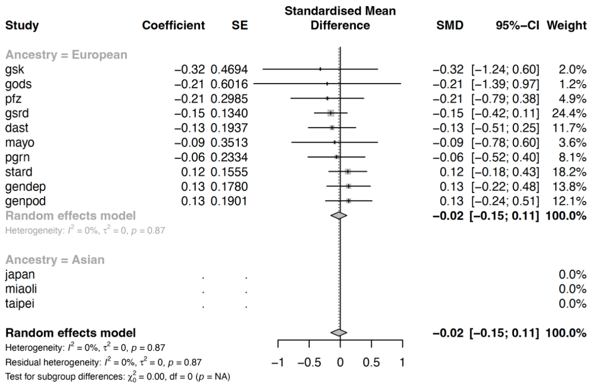

### CYP2D6 intermediate metabolizers

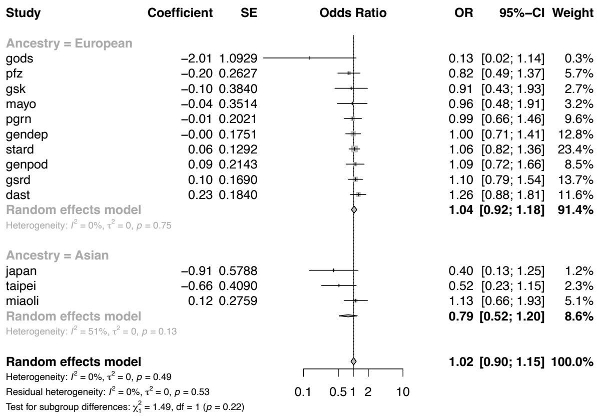

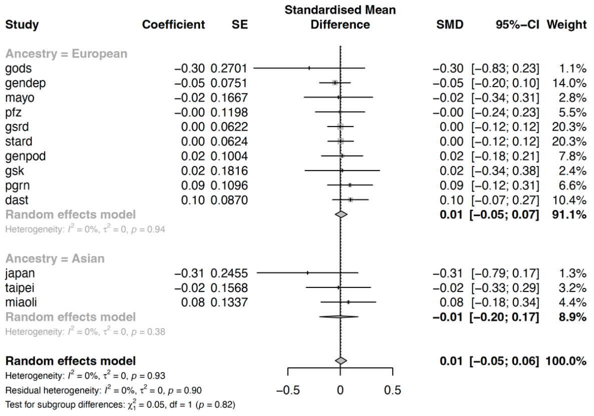

Supplementary Figure 5. Meta-analyses of CYP2C19 poor, intermediate, and ultrarapid metabolizers in CYP2C19 antidepressant group

### CYP2C19 poor metabolizers

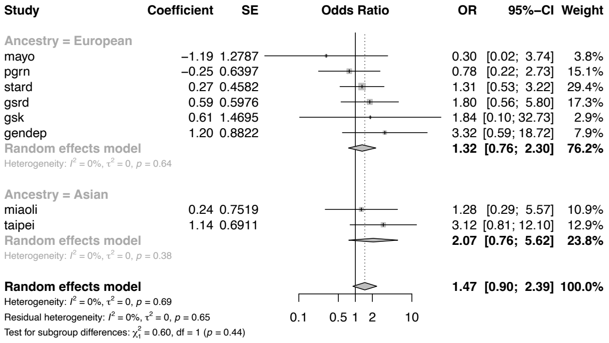

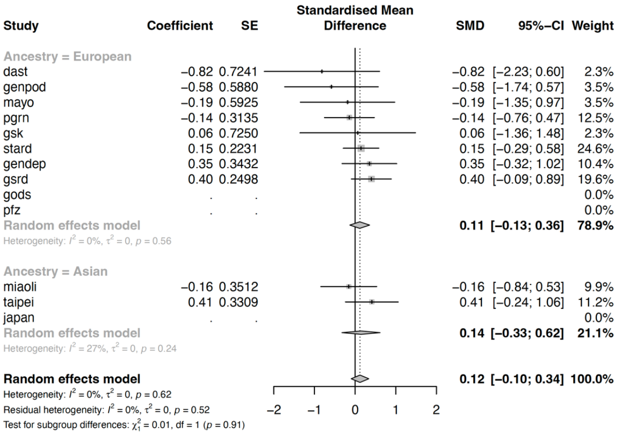

### CYP2C19 intermediate metabolizers

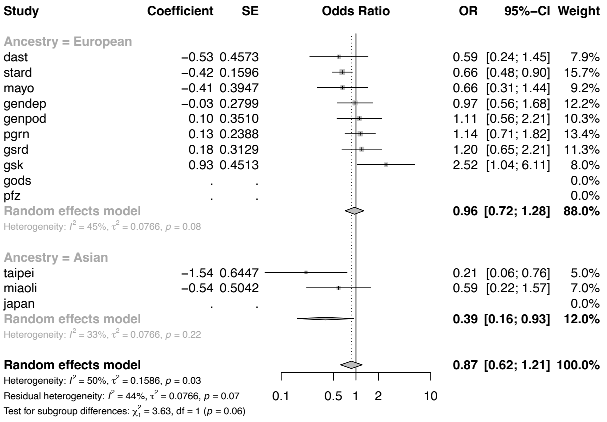

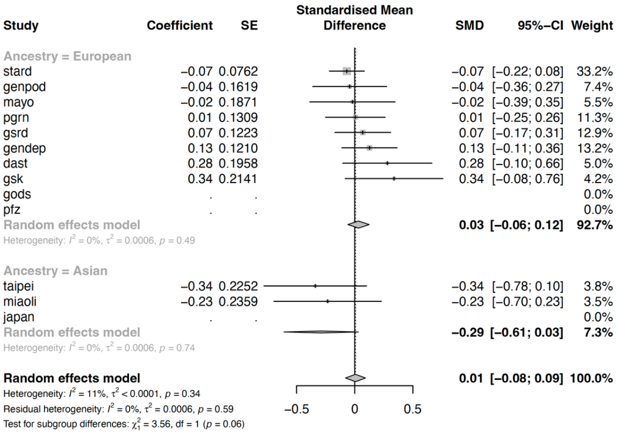

### CYP2C19 ultrarapid metabolizers

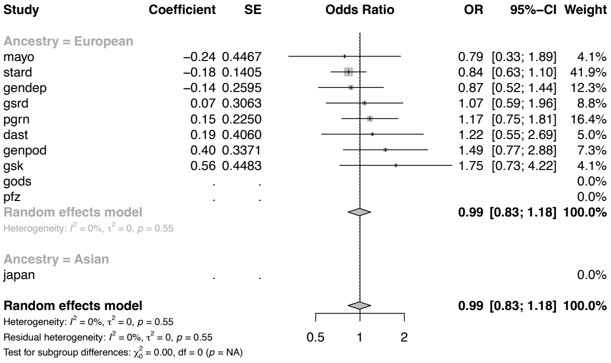

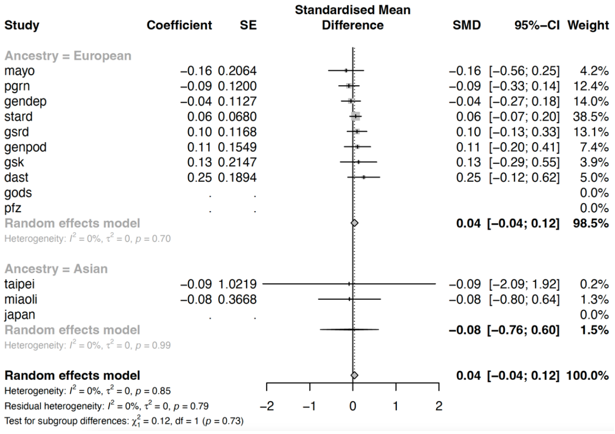

Supplementary Figure 6. Meta-analyses of CYP2D6 poor and intermediate metabolizers in CYP2D6 antidepressant group

### CYP2D6 poor metabolizers

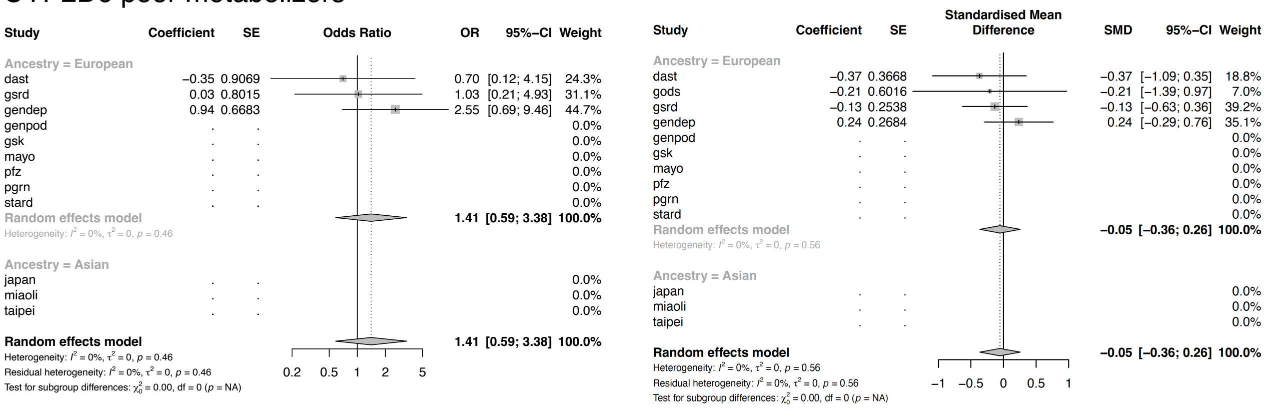

### CYP2D6 intermediate metabolizers

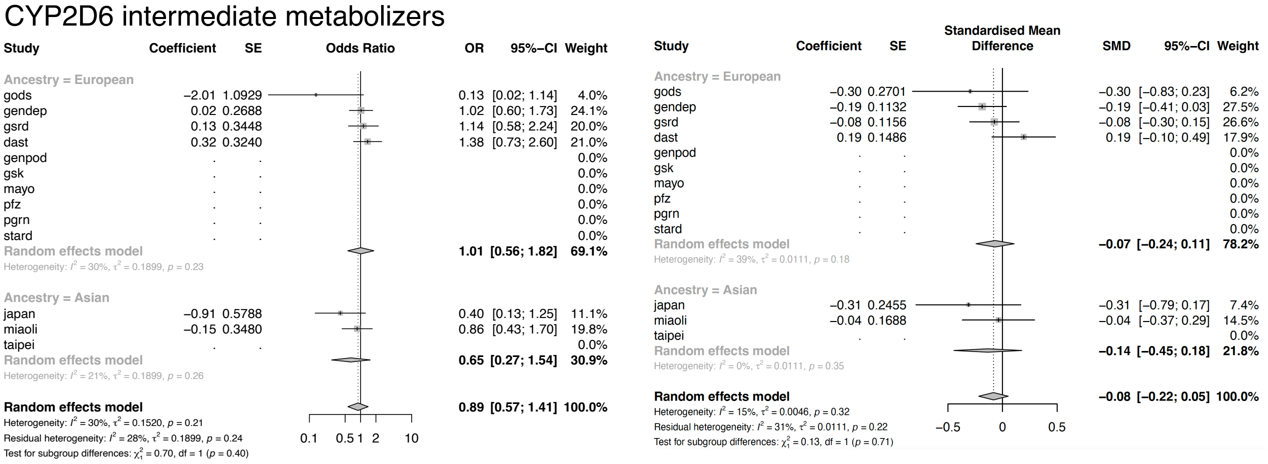

Supplementary Figure 7 Association of metabolizer status with antidepressant outcomes in drugs not primarily metabolised by CYP2C19 or CYP2D6

a. Antidepressants not primarily metabolised by CYP2C19

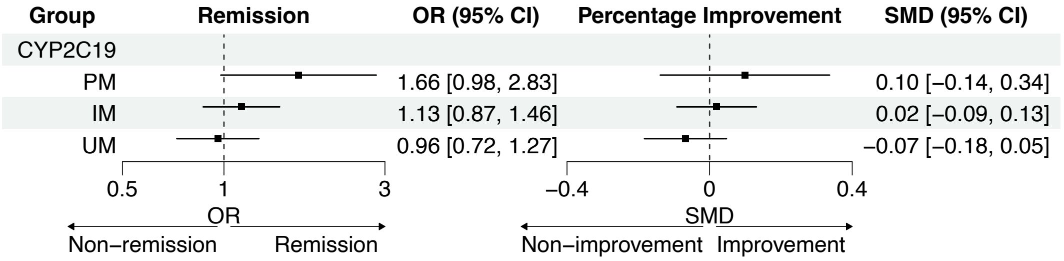

b. Antidepressants not primarily metabolised by CYP2D6

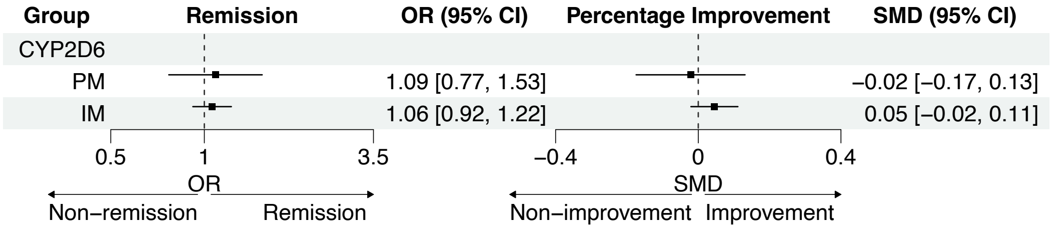

Supplementary Figure 8. Distribution of activity score in each cohort

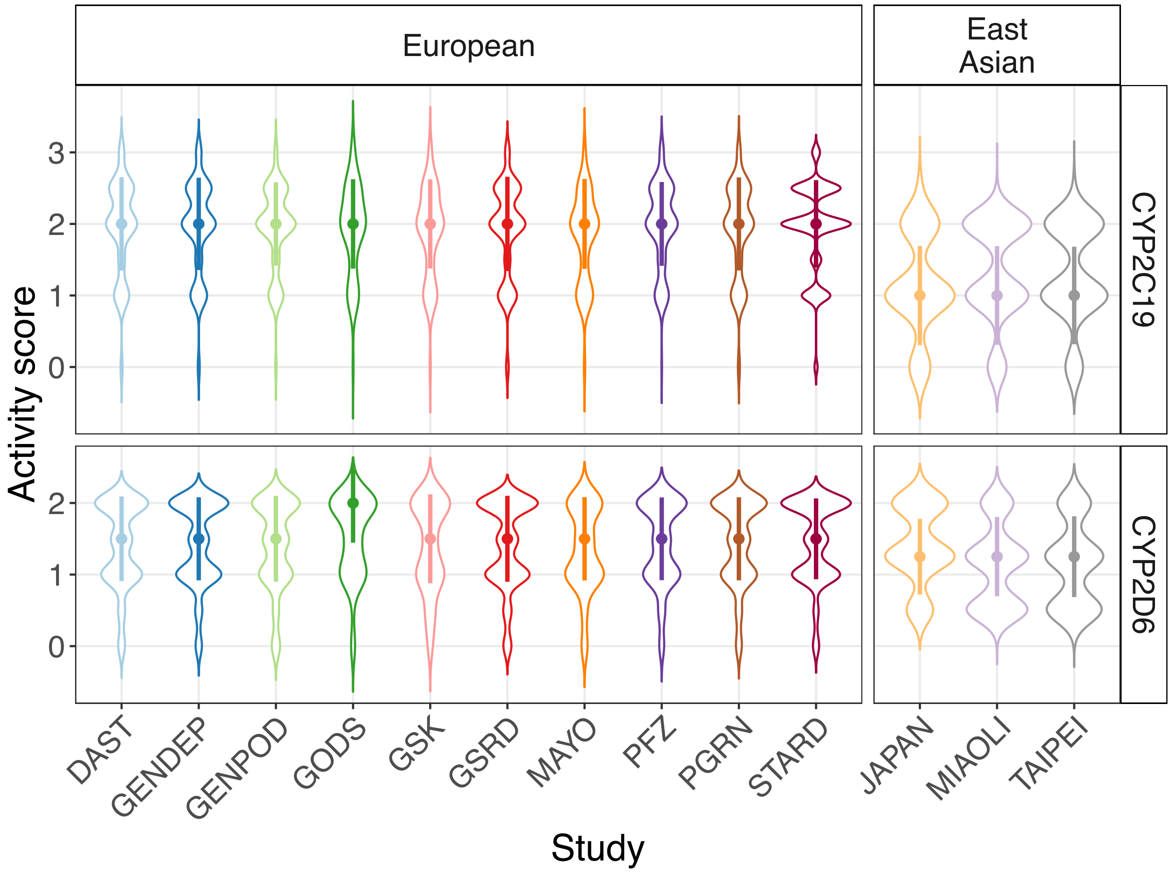

**Supplementary Tables**

Supplementary Table 1. Characteristics of 13 clinical studies

| Studies | Sample size (N = 5843) | Study design | Study weeks | Measures | Antidepressants | Age | Sex (Female) | Remission rate |
|---|---|---|---|---|---|---|---|---|
| **European** | | | | | | | | |
| STARD | 1163 | Open label | 12 | QIDSC | Citalopram | 43.33 (13.49) | 675 (58.0%) | 43.5% |
| GSRD | 1152 | Naturalistic | ≥4 | MADRS | Various | 52.23 (14.02) | 758 (65.8%) | 16.4% |
| GENDEP | 783 | Partially randomized open label | 12 | MADRS | Escitalopram, nortriptyline | 42.28 (11.59) | 490 (62.6%) | 37.2% |
| DAST | 586 | Naturalistic inpatient | 6 | HAMD-21 | Various | 49.47 (15.48) | 335 (57.2%) | 41.8% |
| PGRN | 490 | Open label | 8 | QIDSC | Citalopram, escitalopram | 39.86 (13.64) | 307 (62.7%) | 40.8% |
| GENPOD | 474 | Open label | 12 | BDI | Citalopram, reboxetine | 39.39 (12.50) | 327 (69.0%) | 35.7% |
| PFZ | 309 | RCT | 6-8 | HAMD-17 | Sertraline, fluoxetine, paroxetine | 43.17 (13.06) | 208 (67.3%) | 32.0% |
| MAYO | 156 | Open label | 8 | HAMD-17 | Citalopram, escitalopram | 40.03 (13.88) | 96 (61.5%) | 51.3% |
| GSK | 132 | RCT | 8 | HAMD-17 | Escitalopram | 36.36 (11.90) | 72 (54.5%) | 42.4% |
| GODS | 71 | Open label | 8 | MADRS | Paroxetine | 37.32 (10.34) | 37 (52.1%) | 23.9% |
| **East Asian** | | | | | | | | |
| MIAOLI | 233 | Open label | 8 | HAMD-17 | Escitalopram, paroxetine | 41.36 (13.60) | 192 (82.4%) | 44.2% |
| TAIPEI | 174 | Open label | 8 | HAMD-17 | Fluoxetine, citalopram | 47.01 (15.13) | 96 (55.2%) | 25.9% |
| JAPAN | 120 | RCT | 6 | HAMD-17 | Fluvoxamine, paroxetine | 45.99 (15.25) | 56 (46.7%) | 65.0% |

Mean with standard deviation for age and frequency with proportion for sex were displayed.
BDI, Beck Depression Inventory; HAMD-17, 17-item Hamilton Depression Rating Scale; HAMD-21, 21-item Hamilton Depression Rating Scale; MADRS, Montgomery Åsberg Depression Rating Scale; QIDSC, Quick Inventory of Depressive Symptomatology; RCT: randomized controlled trial.

Supplementary Table 2. Star alleles in CYP2C19 and CYP2D6

| Star alleles | Defining variants | Function | Activity value |
|---|---|---|---|
| CYP2C19 | | | |
| *1 | Reference allele | Normal | 1 |
| *2 | rs4244285 | No | 0 |
| *3 | rs4986893 | No | 0 |
| *8 | rs41291556 | No | 0 |
| *11 | rs58973490 | Normal | 1 |
| *17 | rs12248560 | Increased | 1.5 |
| *35 | rs12769205 | No | 0 |
| CYP2D6 | | | |
| *1 | Reference allele | Normal | 1 |
| *2 | rs16947, rs1135840 | Normal | 1 |
| *3 | rs35742686 | No | 0 |
| *4 | rs3892097 | No | 0 |
| *6 | rs5030655 | No | 0 |
| *9 | rs5030656 | Decreased | 0.5 |
| *10 | rs1135840, rs1065852 | Decreased | 0.25 |
| *14 | rs5030865, rs16947, rs1135840 | Decreased | 0.5 |
| *17 | rs28371706, rs16947, rs1135840 | Decreased | 0.5 |
| *33 | rs28371717 | Normal | 1 |
| *34 | rs16947 | Normal | 1 |
| *35 | rs769258 | Normal | 1 |
| *39 | rs1135840 | Normal | 1 |
| *41 | rs28371725, rs16947, rs1135840 | Decreased | 0.5 |
| *69 | rs28371725 rs1065852 | No | 0 |
| *114 | rs5030865, rs1065852 | No | 0 |

Defining variants were based on the Clinical Pharmacogenetics Implementation Consortium (CPIC) allele definition table.

Supplementary Table 3. Concordance rate of CYP2C19 and CYP2D6 metabolic phenotypes between imputed genotype and Roche AmpliChip CYP450 microarray/TaqMan SNP genotyping in GENDEP

| Imputed genotype | CYP2C19 | CYP2D6 |
|---|---|---|
| Poor | 88.2% | 88.2% |
| Intermediate | 96.1% | 83.8% |
| Normal | 96.6% | 76.7% |
| Ultrarapid | 97.0% | - |

Supplementary Table 4. CYP2C19 antidepressants and CYP2D6 antidepressants

| Gene | Antidepressants |
|---|---|
| CYP2C19 | citalopram, escitalopram, sertraline, amitriptyline, clomipramine, doxepin, trimipramine |
| CYP2D6 | paroxetine, nortriptyline, venlafaxine, fluvoxamine, amitriptyline, clomipramine, trimipramine, desipramine, doxepin |

Supplementary Table 5. Remission and percentage improvement in CYP2C19 and CYP2D6 antidepressant groups

| Metabolizers | N | Remission | Percentage Improvement |
|---|---|---|---|
| CYP2C19 antidepressant group (N = 3390) | | | |
| Poor | 92 | 42 (45.7%) | 0.190 (1.040) |
| Intermediate | 907 | 353 (38.9%) | 0.018 (0.975) |
| Normal | 1343 | 571 (42.5%) | 0.007 (1.037) |
| Ultrarapid | 1048 | 438 (41.8%) | 0.039 (1.016) |
| CYP2D6 antidepressant group (N = 1223) | | | |
| Poor | 43 | 16 (37.2%) | -0.084 (0.892) |
| Intermediate | 433 | 153 (35.3%) | -0.118 (0.973) |
| Normal | 747 | 295 (39.5%) | -0.032 (0.991) |

Frequency with proportion for remission and mean with standard deviation for percentage improvement were displayed

Supplementary Table 6. Meta-analyses of activity score with antidepressant outcomes

| Outcomes | OR/COR | 95% CI | P |
|---|---|---|---|
| CYP2C19 activity score | | | |
| Remission | 0.94 | 0.86, 1.03 | 0.197 |
| Percentage Improvement | -0.02 | -0.05, 0.01 | 0.170 |
| CYP2D6 activity score | | | |
| Remission | 1.01 | 0.90, 1.14 | 0.882 |
| Percentage Improvement | 0.001 | -0.03, 0.03 | 0.958 |

COR: correlation

Supplementary Table 7. Meta-analyses of metabolic phenotypes for percentage improvement adjusting for baseline severity of depression

| Metabolizers | SMD | 95% CI | P |
|---|---|---|---|
| CYP2C19 | | | |
| Poor | 0.13 | -0.03, 0.29 | 0.103 |
| Intermediate | 0.01 | -0.05, 0.08 | 0.683 |
| Ultrarapid | 0.01 | -0.05, 0.07 | 0.777 |
| CYP2D6 | | | |
| Poor | -0.02 | -0.15, 0.11 | 0.712 |
| Intermediate | 0.01 | -0.05, 0.07 | 0.724 |
